# CancerSpot: A multi-cancer early detection test developed and validated on a retrospective cohort

**DOI:** 10.1101/2024.12.03.24318395

**Authors:** Swaraj Basu, Prakash Hiremath M, Nihesh Rathod, Aditi Chatterjee, Divya Vishwanath, Arunima Ghosh, Sweta Sanguri, Sampuran Chakraborty, Aastha Tripathi, RT Preetha, Arya Nair, Goutham Kumar, Kannadhasan Sekar, Subuhi Yete, G Bhanumathy, Urvashi Bahadur, Aneesha Radhakrishnan, Ankita Khan, Yasodha Kannan S, Lavanya Bollipalli, Pallavi Ghana, Aparnaa Ramanathan, Puja Saha, Sameer Phalke, Charles Cantor, Sewanti Limaye, Vijay Chandru, Vamsi Veeramachaneni, Ramesh Hariharan

## Abstract

Next-generation sequencing (NGS) technologies have transformed biomarker discovery, enabling the detection of disease-associated markers at the earliest stages of illness. In this study, we introduce a blood-based, non-invasive test for multi-cancer detection using cell-free DNA (cfDNA) methylation sequencing. The test employs a novel methylation scoring system derived from sequencing data and integrates machine learning to analyze a retrospective cohort of newly diagnosed cancer cases and controls recruited from multiple centers across India. To enhance robustness, the study includes a substantial proportion of controls with habitual tobacco and alcohol use, ensuring the test’s resilience against confounding factors. The test’s accuracy was further validated through synthetic data augmentation, demonstrating reliability under conditions of random signal perturbation. At an approximate specificity of 97%, the assay achieves sensitivities of 79.3% for Stage I, 78.4% for Stage II, 78.4% for Stage III, and 86.8% for Stage IV cancers in an independent validation cohort. Additionally, the test demonstrates Top 2 Tissue of Origin (TOO) accuracies of 78.3% for Stage I, 79.3% for Stage II, 82.8% for Stage III, and 69.7% for Stage IV cancers. This blood-based test holds considerable promise for early cancer detection, offering a precise test for cancer screening.

## Introduction

Cancer accounts for a staggering 1 in 6 deaths globally (16.8%), according to recent estimates where lung, breast, colorectal, prostate, stomach, and liver cancers collectively contribute to nearly 50% of cancer incidence and mortality worldwide [1]. In the United States alone, approximately 2 million new cancer cases are projected for 2024 [2] while in India around 1.4 million cases were estimated for the year 2022 which is expected to rise at a rate of 12.8% by 2025 [3]. A recent study has unequivocally demonstrated that the “cure fraction” - defined as the proportion of cancer patients who survive beyond the early peak in cancer-specific mortality - is significantly higher for cancers detected at an early stage [4]. For select cancers, the study highlights the dramatic decline in the cure fraction from Stage I to Stage IV: **Breast Cancer** (88%, 82%, 59%, 6%), **Lung Cancer** (74%, 47%, 21%, 5%), **Colorectal Cancer** (62%, 61%, 58%, 7%), **Esophageal Cancer** (60%, 43%, 27%, 5%), and **Liver Cancer** (32%, 32%, 8%, 3%), thus providing an ample evidence in support of better treatment outcomes if cancer is diagnosed at an early stage.

Liquid biopsy-based cancer detection and screening offer significant promise due to their non-invasive nature and ability to detect multiple cancer types within a single test [5]. This approach relies on analyzing cell-free DNA (cfDNA) fragments released into the bloodstream through active secretion or cell death [6] where DNA methylation patterns, variations in cfDNA fragment size, fragment end-point motifs, or mutations can help distinguish tumor-derived cfDNA from background cfDNA originating from normal cells [7]; [8]. DNA methylation signatures, in particular, have shown remarkable accuracy in identifying specific cancer types, such as colorectal cancer [9] and lung cancer [10], with sensitivities exceeding 80% and 90%, respectively at a specificity greater than 90%. Beyond these select cancer types, multi-cancer early detection (MCED) tests using liquid biopsies have gained traction leading to several studies reporting over 90% accuracy across a wide range of cancers, particularly in the early stages [11];[12];[13]. Despite these advancements, a significant challenge in cfDNA-based cancer diagnostics lies in accurately identifying the small fraction of tumor-derived cfDNA amidst the high background levels of normal cfDNA in the bloodstream [14];[15].

A key focus in developing a MCED platform via liquid biopsy is creating a cost-effective assay that maintains high performance. This can be achieved through targeted sequencing [16], where markers of interest are identified from published literature, public datasets [17], or in-house sample cohorts [11];[12]. In this context, we introduce CancerSpot, the first MCED test based on targeted methylation sequencing across a 123 Mb panel developed using a diverse Indian population cohort. The test utilizes enzymatic methylation sequencing (EM-seq), which offers significantly lower DNA damage and reduced GC bias compared to conventional bisulfite methylation sequencing assays [18]. The study analyzes cfDNA derived from blood plasma across a cohort of approximately 688 cancer patients (spanning 10 cancer types) and 326 controls to develop a robust genomics pipeline on which a machine learning framework was tuned for cancer classification and tumor type prediction to identify ∼17000 methylation markers. CancerSpot demonstrates a sensitivity of 78% for detecting Stage I cancers (at ∼97% specificity) and achieves 77% accuracy in identifying the correct tissue of origin within the top two predicted tumor types. Additionally, we also identified markers that were derived from an extensive analysis of publicly available methylation datasets and a thorough curation of relevant literature. De novo markers identified from our sample cohort demonstrated superior performance compared to markers that can be obtained from public data sets and literature, despite a low degree of overlap. This underscores both the utility of mining public databases and the added value of analyzing unique sample cohorts to identify robust and clinically relevant markers.

## Methods and Materials

### Clinical sample collection

This multicentric study involved four distinct cohorts recruited across various hospital centers in India (Supplementary Table 1). The study protocol was approved by the ethics committees for each participating center, ensuring compliance with ethical guidelines (CTRI No: CTRI2022/05/042936). Participation was voluntary, and all subjects provided explicit informed consent prior to enrollment. Cohorts 1, 2, and 3 included treatment-naive patients aged 18 years or older who met the following criteria: a) no prior oncological treatment or surgery, b) diagnosed or suspected to have benign, precancerous, or malignant neoplasms and/or lesions, and c) deemed eligible for onco-surgery or oncological treatment, as determined by the site’s Principal Investigator (treating oncologist) or Co-Investigators through routine clinical screening processes. Patients with recurrent or relapsed cases were excluded. Anyone who had undergone prior onco-surgery or oncotherapy were excluded. Cohort 4 consisted of individuals aged 50 years or older who were seemingly healthy with no symptoms or suspicion of cancer, with no history of benign, precancerous, or malignant neoplasms and/or lesions, and no prior cancer diagnoses. Recruitment was conducted by the site’s Principal Investigator or Co-Investigators through routine clinical care or screening procedures. Subjects across all cohorts were stratified based on lifestyle habits, including tobacco use (chewers only, smokers only, those with mixed habits) and moderate habitual alcohol consumption, as well as those without these habits. Detailed recruitment criteria and stratification methods are provided in the Supplementary Document.

A total of 3396 subjects with benign, precancerous or malignant neoplasms and/or lesions with primaries in the breast, stomach, esophagus, ovary, cervix, lung, colorectal, pancreas, liver, and gallbladder were recruited from 39 sites along with 1181 controls. Cases of uncontrolled infection were excluded, as were pregnant or lactating women. Subjects who were coagulopathic and taking blood thinning products were included based on the site PI’s assessment on safety for a blood draw. Whole blood was collected in 2x10 ml plasma preparation tubes with anticoagulants (Streck Tubes) and gently mixed. Matched FFPE blocks (treatment naive) from malignant and benign lesions were collected wherever available. Samples were shipped at room temperature and reached the Strand Life Sciences reference laboratory in Bengaluru, India within 48-72 hours.

### Methylation assay and sequencing

A subset of 1,016 cancer samples and 376 control samples was selected for downstream analysis using a targeted MethylSeq assay. Blood samples collected in Streck tubes were centrifuged at 1900 × g for 22 minutes at 4°C. The supernatant was carefully transferred to 15 mL tubes and subjected to a second centrifugation at 3200 × g for 20 minutes at 4°C. The resulting plasma was manually extracted and stored in 4 mL aliquots at -80°C. The buffy coat was separated, preserved in RNAlater, and stored at -80°C for further analysis. Cell-free DNA (cfDNA) was extracted from 4 mL of plasma using the Apostle MiniMax High Efficiency cfDNA Isolation Kit (Cat. No.: A17622-384) following the manufacturer’s protocol (Apostle Bio, <u>link</u>). The extracted cfDNA was quantified using a Qubit High Sensitivity dsDNA kit (Q32854, Invitrogen), and its size distribution was evaluated using the Agilent 4200 TapeStation (<u>link</u>) with High Sensitivity DNA1000 ScreenTapes (<u>link</u>). Typical cfDNA profiles on the TapeStation revealed a prominent peak at ∼190 bp and a secondary peak in the 500–700 bp range. Samples with at least 10ng of cfDNA were taken forward for library preparation.

MethylSeq libraries were prepared from extracted cfDNA using the NEBNext Enzymatic Methyl-Seq Kit (<u>link</u>) following the manufacturer’s protocol. In brief, 10–20 ng of cfDNA, spiked with 0.02ng of unmethylated lambda and 0.001ng of methylated pUC19 was subjected to end-repair, A-tailing, and adapter ligation. TET2 and T4-BGT enzymes were employed to protect 5mC and 5hmC from deamination. A bead-based cleanup step was performed using the purification beads provided with the kit, followed by a safe stop step at -20°C, maintained for at least 48 hours. This pause was critical for ensuring high conversion rates during subsequent steps. Unprotected cytosines were converted to uracils using APOBEC3A. Libraries were then PCR amplified, with the number of cycles tailored to the DNA input amount (e.g., 11 cycles for 10–14 ng, 10 cycles for 15–20 ng). Unique barcoded indices were introduced during the PCR step. The libraries were quantified using a Qubit High Sensitivity dsDNA Kit, with a yield of ≥400 ng considered acceptable. Size distribution was assessed using the Agilent 4200 TapeStation with DNA1000 ScreenTapes. Typical cfDNA libraries displayed a prominent peak around 350 bp and a secondary peak near 500 bp. A library with a yield of 400 ng or more was subsequently taken ahead for capture.

A quantitative PCR method was used to estimate the efficiency of conversion of unmethylated cytosine to uracil. This method used primers that amplify selected regions of the highly repetitive Alu sequences in the human genome. The Alu primer binding sites contain multiple cytosines. In fully converted samples, these cytosines would be converted to uracil and reduce the binding efficiency of the primers, thereby requiring a large number of PCR cycles for amplification. On the contrary, if conversion is not complete, many regions would still resemble the original primer binding regions, enabling amplification within the first few cycles of PCR. Libraries that took at least 21 cycles for amplification were considered to have undergone complete conversion, and were taken ahead for hybridisation with target probes (details in Supplementary Document).

Hybridisation capture of the target regions was performed overnight with the Twist Human Methylome panel probes (<u>link</u>) comprising 551,803 target regions with 123 million target bases covering 3.98 million CpGs, by pooling 187.5 ng of eight MethylSeq libraries with distinct indices. The captured regions were amplified using 7 cycles of PCR, generating a post-capture library. This library was quantified using the Qubit High Sensitivity dsDNA kit, and size estimation was carried out by running it on the TapeStation 4200 using the DNA1000 tape. A typical post-capture library has a concentration of around 25-35 ng/μL and has an average size of around 350bp. Libraries were sequenced at 2x150 bp reads (paired-end sequencing) either on the Illumina NovaSeq 6000 (S4 flowcells) or on the Illumina NovaSeq X Plus (10B/25B flowcells), aiming for 45 Gb of data.

### Methylation data analysis pipeline

Demultiplexing was conducted using **bcl-convert** version 1.0.0 (<u>Illumina</u>), producing FASTQ files that were subsequently processed through our in-house pipeline, **FrAnaTk** (Fragment Analysis Toolkit). Workflow management in FrAnaTk was handled using **Snakemake** version 7.17.1 [19]. **BbDuk** version 38.96 (<u>SourceForge</u>) was employed for adapter removal and low-quality read trimming with the following parameters: k=19, mink=5, hdist=1, hdist2=0, ktrim=r, qtrim=r, minlength=36, and trimq=14. Read quality assessment was performed using **FASTQC** version 0.12.1 (<u>Babraham Bioinformatics</u>). Reads were aligned using **BWAMeth** version 0.2.5 [20], which accommodates APOBEC-induced deamination of unmethylated cytosines by aligning reads to two versions of a custom reference genome (hg38 - UCSC; Lambda - J02459.1, pUC19 - L09137.2, HPV16 - NC_001526.4, HPV18 - NC_001357): one with intact cytosines and the other with cytosines converted to thymines. Duplicate reads were marked using **Samblaster** version 0.1.26; [21], and alignment files were sorted and indexed with **samtools** version 1.14 [22]. For methylation analysis, the sorted BAM files were processed with **MethylDackel** version 0.6.1 (<u>GitHub</u>), generating bedGraph files that detail cytosine and thymine read counts for each CpG, CHG, and CHH site. **MethylDackel extract** was first used to create basic bedGraphs, followed by **MethylDackel mergecontext** to annotate the proper start and end positions for each context. To compute cfDNA fragment length, end-motifs, and read-wise methylation metrics, additional filtering was performed using **samtools** with the parameters -f 3 -F 3852 -G 48. This retained only properly paired reads, excluding unaligned reads, PCR or optical duplicates, supplementary alignments, and reads failing quality control. Filtered BAM files were converted into fragment bed files using **bedtools** version 2.30.0 [23] following a methodology adapted from **FinaleDB**, a published fragmentomics database [24]. The outputs from these steps were integrated to generate various quality control (QC) metrics using a combination of tools, including **Samtools** , **bedtools** , **R** version 4.1.12, **Picard** version 2.18.29 (<u>GitHub</u>), and **Python**.

### Derivation of metrics for downstream analysis

A customized version of **wgbs-tools** [25] was utilized to generate a **PATR** file from the BAM file. By default, **wgbs-tools** creates CpG-clustered PAT files; however, the customized version was configured to produce a PATR file that merges both mates of a read pair into a single fragment, excludes fragments that do not overlap any CpG sites, and organizes the remaining fragments one per line. Each line in the PATR file represents a fragment and indicates, for each overlapping CpG, whether the fragment contains a cytosine (C) or a thymine (T). Filters were applied during PATR file generation to select reads based on the following criteria:

● **SAM flag filters**: Reported above (-f 3 -F 3852).
● **Mapping quality**: Reads with a mapping quality score ≥ 30 were retained.

The conversion efficiency of each read was calculated using a novel, orientation-agnostic method. Reads were assessed based on their conversion efficiency, determined from non-CpG cytosines (Cs) and guanines (Gs) in the reference genome ((hg38 - UCSC; Lambda - J02459.1, pUC19 - L09137.2, HPV16 - NC_001526.4, HPV18 - NC_001357)). Only reads with a conversion efficiency ≥ 95% were included in downstream analyses. Details of this approach are provided in the Supplementary Document. The **Bedgraph** and **PATR** files were further analyzed to derive various metrics for each sample:

● **Percentage conversion (pConvNonCpG)**: Calculated as the total number of reads with thymine
(T) substitutions across all CHG and CHH sites, divided by the total number of reads across these sites.
● **Percentage conversion using lambda control (pConvCpGLambda)**: For assessing the efficiency of enzymatic conversion by comparing the methylation status of unmethylated lambda DNA to ensure accurate detection of methylated cytosines.
● **Average coverage (cov.avg)**: Derived from the **hsmetrics** output file generated by **Picard CollectHsMetrics**, which includes coverage statistics and the number of high-quality bases for each coverage level.
● **On target alignment rate (OnTrgtAln)**: The fraction of reads which are aligned within the targets defined by the 123 Mb Twist methylome panel reflecting the efficiency of capture.
● **Percentage of duplicate reads (pDup)**: Fraction of reads marked as duplicates indicating the DNA quality, PCR amplification and library complexity.

**Fragment-Wise Methylation Scores** were calculated for each region in the targeted panel (551,803 regions) using the **PATR file** for each sample. For each region, the following metrics were computed:

● We identified **useful fragments** defined as, the fragments with at least three CpGs with at least one CpG falling within the target region.
● Among these useful fragments, the proportion with extreme methylation fractions was calculated:

○ **Highly methylated fragment fraction (HMF or Ge80)**: Fraction of fragments with ≥80% of CpGs methylated.
○ **Poorly methylated fragment fraction (LMF or Lt20)**: Fraction of fragments with <20% of CpGs methylated.

**Global Fragmentomic Scores** were derived from the fragment bed file (generated from the BAM file) for each sample. Two sets of 256 scores were computed, corresponding to the 256 possible 4-mer sequences along with fragment length profile [26]:

1. **Endpoint scores (E-AAAA, etc.)**: Calculated as the fraction of fragments overlapping the target regions where the 5’ end of the fragment matches the given 4-mer in the reference sequence (hg38 - UCSC).
2. **Breakpoint scores (B-AAAA, etc.)**: Determined as the fraction of fragments overlapping the target regions where the 5’ end of the fragment aligns precisely in the middle of the given 4-mer in the reference sequence (hg38 - UCSC).
3. **Fragment length profile**: Fraction of fragments falling into a particular bin defined for size (for eg. <120 Bp, 120-150 Bp, 150-170 bp etc)

### Analysis of publicly available methylation datasets

Data from the Illumina HumanMethylation 450K BeadChip, covering approximately 485,547 CpG sites across the human genome, was compiled from multiple sources:

● The Cancer Genome Atlas (<u>TCGA</u>): Provides standardized data on cancer tissues and adjacent normal tissues across various cancer types.
● Epigenome-Wide Association Study (EWAS) (<u>datahub</u>): Includes a subset of TCGA and Gene Expression Omnibus (GEO) datasets processed using a unified pipeline. EWAS also incorporates data on healthy tissues and blood samples.
● Gene Expression Omnibus (GEO) (<u>source</u>): Offers a broad range of datasets generated using diverse treatments and normalization methods, which may introduce variability.

A total of 4,475 tumor tissues and 393 adjacent normal tissues from TCGA, 1,789 normal tissues and 600 blood samples from EWAS, and 656 blood samples from GEO were compiled. From this dataset, differentially methylated seed probes across multiple cancer types were identified and extended into hyper-methylated and hypo-methylated differentially methylated regions (DMRs), as detailed in the Supplementary Document.

### Curation of literature for methylation markers

To further enhance the analysis, 391 publications were curated for probes, genes, and CpG sites reported as hyper- or hypo-methylated in cancer versus normal samples. This curation followed a two-step strategy:

1. Initial Search: PubMed was queried using keywords focusing specifically on cfDNA based early detection studies (e.g., “methylation,” “cancer,” “cfDNA,” “early diagnosis”), resulting in approximately 100 PMIDs. While cancers like colorectal, lung, and breast cancer were well represented, other cancers of interest in the Indian population, such as head and neck, cervical, esophageal, and gastric cancers, were underrepresented.
2. Revised Search: Relaxing the keyword constraints to include cancer-specific queries and cancer specific methylation signals (from non cfDNA or early detection studies as well) for nine cancers of interest yielded an additional 290 PMIDs, bringing the total to 391 curated PMIDs.

These publications primarily employed three approaches:

1. Sequencing/array-based techniques (e.g., whole-genome bisulfite sequencing (WGBS), microarrays).
2. Analysis of public datasets, including TCGA and GEO.
3. Predetermined targets selected through literature reviews, often analyzed using PCR-based methods.
4. Markers identified are detailed in Supplementary Table 5.

### Cancer Detection and Tissue of Origin models

Model development incorporated the **Ge80 score** described earlier, alongside the yield in ng per ml of plasma, as key features. The data was split into two sets: a) **Leave-out set (20%)**: Randomly selected 20% of samples from each category, reserved for independent testing b) **Leave-in set (80%)**: Used for model training and evaluation through 4-fold cross-validation. **Stratification** during the train-test split ensured balanced representation based on **cancer type, stage, and gender**. The binary classificationML workflow involved **categorized** samples into two classes: **class0** (e.g., control) and **class1** (e.g., cancer) followed by

1. **Cross-Validation**:

○ The leave-in set was split into 4 folds, and cross-validation was performed **20 times**, resulting in **80 models** (20 × 4).
○ The validation process operated solely on the leave-in set, without accessing the leave-out set.
2. **Threshold Determination**: A threshold for classification was defined using the predicted scores of **class0** samples from the validation folds of the leave-in set, based on a desired specificity.
3. **Leave-Out Set Evaluation**:

○ The **80 models** were applied to the leave-out set.
○ Each sample in the leave-out set was assigned an **average prediction score** across the models.
○ Samples were classified as **class1** if the score exceeded the threshold; otherwise, they were assigned to **class0**.

Region selection was an integral step in the cross-validation process. It was performed on the **training subset of the leave-in set** before model building with **Mann-Whitney U test** to rank regions by significance and selection of 1000 significant regions per fold (250 regions per fold for TOO models) were selected as features for model building. **XGBoost** (<u>documentation</u>), a non-parametric ensemble learning method based on decision trees was used for the classification (supervised learning). The **reg_alpha** parameter was optimized during training, while other parameters were kept at their default settings.

To improve robustness against technical variability, **synthetic sample augmentation** was applied for each sample in the leave-in set, where **10 perturbed copies** were generated. Perturbations were derived from the observed distribution of **Ge80 differences** of a pool of samples run on two sequencers (**NovaSeq 6000** and **NovaSeq XPlus**). Combined with the original sample, this resulted in **11 copies per sample** used for training. This strategy ensured the model was robust to feature variations and reduced the likelihood of overfitting to specific technical artifacts. **Benign samples** were also included as controls during training to minimize the misclassification of benign samples as cancer in the leave-out set. Further, to evaluate whether the model was learning true cancer-versus-control differences, an additional analysis was conducted by building models using only **covariates** such as age, gender, tobacco usage, and comorbidities. This analysis helped determine if these variables alone were predictive, ensuring that the primary model relied on true biological signals rather than confounding factors.

The cancer types analyzed in this study included colorectal, esophagus, gallbladder, liver, lung, pancreas, stomach, breast, cervix, and ovary. The modeling approach involved a hierarchical schema with two main sets of models: one for cancer detection (L1) and another for tissue of origin (TOO) identification (L2), alongside a standalone model set designed for specific cancer classifications. The standalone models were not used further in the classification, but the regions are in majority common with grouped cancer models, and hence retained for creation of a smaller targeted panel downstream. The L1 model set was designed to determine the cancer status of a sample. It comprised two models: **GenCncrs** and **FmlCncrs**. The **GenCncrs** model used controls as class0 and gender-agnostic cancers (colorectal, esophagus, gallbladder, liver, lung, pancreas, and stomach) as class1. The **FmlCncrs** model used female controls as class0 and female-specific cancers (breast, cervix, and ovary) as class1. Thresholds for these models were set based on desired specificity within the leave-in dataset. For male samples, cancer status was determined solely using the **GenCncrs** model. Female samples were evaluated with both models, and a cancer-positive status was assigned if either model classified the sample as cancer-positive. Thresholds for these models were calibrated to achieve >95% specificity (considering class0) within the leave-in dataset.

The L2 model set was designed for tissue of origin (TOO) determination and included ten cancer-specific models. Each model was binary, with one specific cancer type as class0 and the remaining cancers as class1 (for example **CrcVsRestCncr** for Colorectal cancer vs rest). For male samples, predictions were derived from models excluding female-specific cancers (**BrsVsRestCncr**, **CvxVsRestCncr**, and **OvrVsRestCncr**). Female samples were evaluated with all ten models. The prediction scores from these models were compared to their respective thresholds, and delta values (prediction score minus threshold) were calculated. The cancer types corresponding to the three lowest delta values were used to identify the top three likely tissues of origin. The TOO evaluation was conducted only for samples identified as cancer-positive by the L1 model set.

In summary the leave-in/training set included 261 controls and 532 cancer samples, distributed as follows: Benign (43), Stage I (102), Stage II (125), Stage III (133), and Stage IV (129) while the **leave-out** validation set consisted of 65 controls and 156 cancer samples: Benign (15), Stage I (29), Stage II (37), Stage III (37), and Stage IV (38). Detailed distributions of sample characteristics, including gender, age, and lifestyle habits across cancer stages, are provided in Figure 3 (Supplementary Table 3). Fragment coverage across approximately 552K genomic regions was assessed in the leave-in control samples. Regions with at least 30x coverage in at least 50% of the samples were designated as **coverage-pass** regions, totaling ∼277,000 regions. The initial model training was conducted using Ge80 data from these regions with further refinement by selecting significant regions based on fold change between cancer vs control groups (**Mann-Whitney U test**) focusing on top-ranking regions for each model. Control vs. Cancer Group models were developed using region-wise Ge80 features from the selected regions, alongside cfDNA yield (ng/mL plasma) as a sample-level feature. Model thresholds were calibrated to achieve 95% specificity within the leave-in set. Tissue of Origin (TOO) models were similarly built using region-wise Ge80 features, with thresholds optimized for 91% sensitivity within the corresponding leave-in class 0 samples. Importantly, samples were evaluated for TOO classification only if they were first detected as cancer-positive. The combined union of all selected regions across models amounted to 16,937 regions. Annotation of the regions was performed by the following

- Promoters: UCSC refSeq genes track (+1500, -500 from TSS): <u>link</u>
- Enhancers: Conserved regulatory elements track: <u>link</u>
- Exon/Intron: Using the csaw [27] package in R

Both the L1 and L2 model exercise was also repeated for regions derived from TCGA or from curation by mapping those regions (hypermethylated DMRs from TCGA or hypermethylated promoter/probe coordinates from curation) to the Twist methylome panel, selecting the subset and repeating the ML exercise (including the Mann-WhitneynU test). The same process was performed with random regions, drawn from the entire Twist methylome panel (count matched to the regions selected in L1 and L2 models).

## Results

### Sample collection

Blood samples from patients and controls were collected from 39 centers across India, ensuring a diverse cohort and accounting for natural variability in collection and transport conditions. Notably, 90% of the samples were transported to the Strand laboratory in Bengaluru, India and processed for plasma extraction within 72 hours. The complete workflow—from sample collection through analysis—along with the frequency and distribution of samples across sites, is depicted in Figure 1. A detailed breakdown of sample collection by site and the criteria used for patient staging can be found in Supplementary Table 1. Samples were collected in 1 or 2 Streck tubes (10 mL each), based on individual consent and health status. Importantly, no bias was observed in the volume of blood collected with respect to age (t-test, *P=0.7526*) however a higher fraction of samples had a single tube of blood for late stage patients (25%) compared to early stage (18%, fisher-test, *P=1.59e-6*) (Supplementary Figure 1).

**Figure 1:**
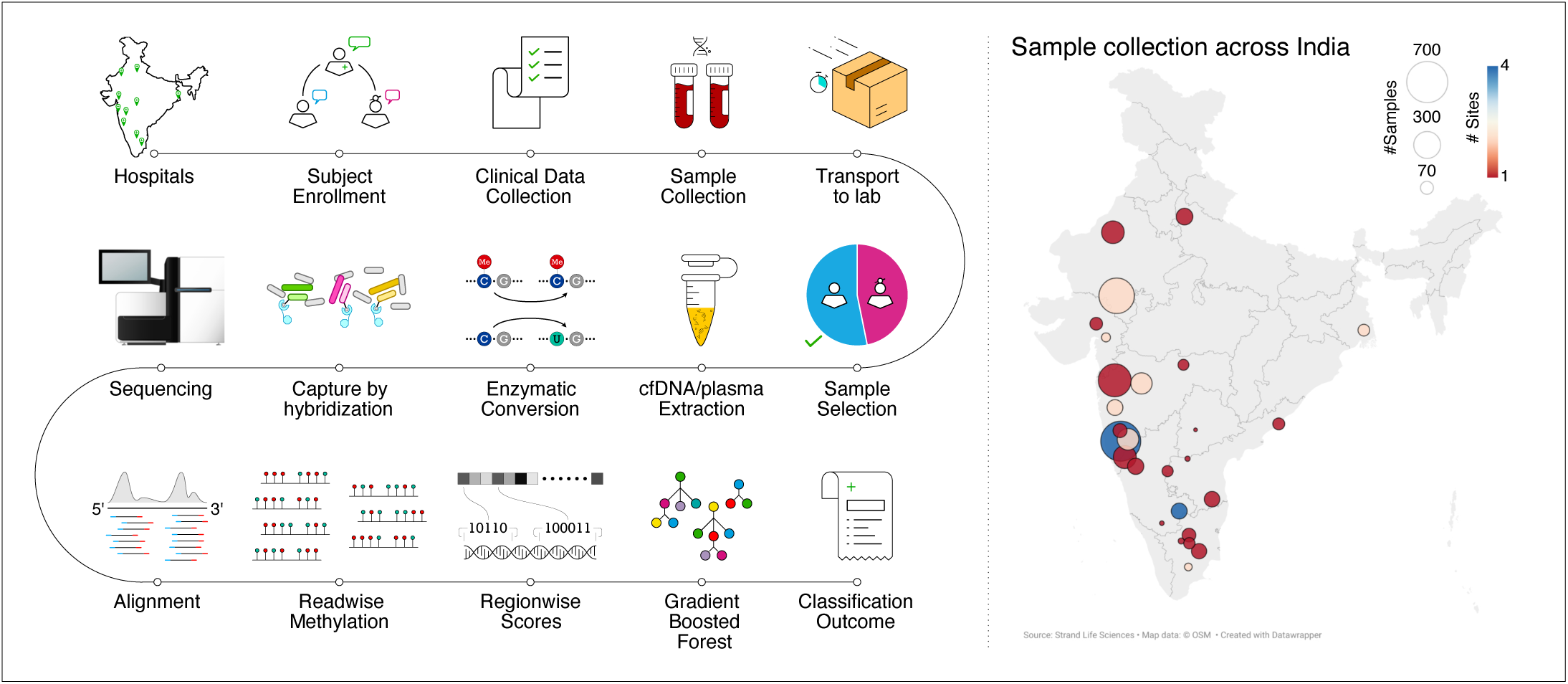
Workflow of sample collection and analysis. This figure provides an overview of the workflow, beginning with sample collection and selection, followed by cfDNA extraction, enzymatic conversion, hybridization-based capture, sequencing, alignment, and data analysis. The workflow culminates in the classification of each sample as either control or cancer. Additionally, the distribution and frequency of samples collected across various sites is shown, highlighting the geographic diversity and representativeness of the dataset. This visual summary underscores the systematic approach and scope of the study design.

### Methylation assay and analysis

Samples meeting the quality threshold of average coverage >= 30x across the 123 Mb panel were included which resulted in a final cohort of 688 cancer samples and 326 controls. The cancer cohort encompassed patients with colorectal, esophageal, gallbladder, liver, lung, pancreatic, stomach, breast, cervical, and ovarian cancers. The analysis workflow involved several stages: sample collection, selection for downstream processing, and machine learning-based modeling. Models were trained on a “leave-in” dataset and evaluated on a separate “leave-out” dataset, as illustrated in Figure 2. Cohort characteristics, including age, gender, body mass index (BMI), tobacco use, alcohol consumption, and comorbidities such as hypertension and diabetes, are summarized in Figure 3 and Supplementary Table 2 (also provides detailed information on tumor site distribution, sample-specific data, and QC metrics obtained from custom methylation and fragmentomics analyses).

**Figure 2:**
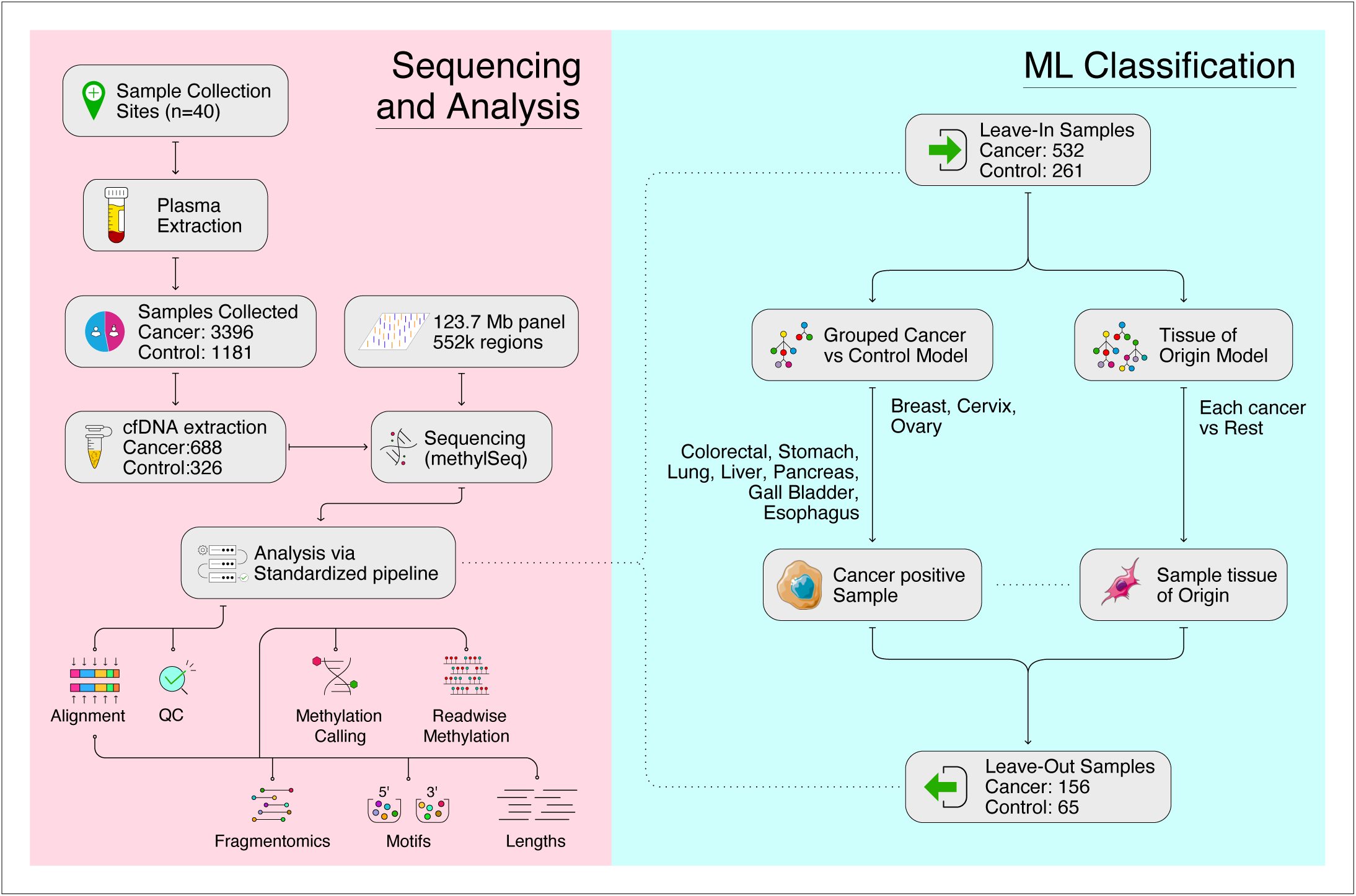
Detailed Workflow of Sequencing, Analysis, and Machine Learning Classification. Detailed breakdown of the workflow, which is divided into two key stages: Sequencing and Analysis, and Machine Learning (ML) Classification. In the **Sequencing and Analysis** stage, the workflow begins with sample collection, followed by plasma and cfDNA extraction. The extracted cfDNA is then sequenced using a 123.7Mb panel that covers 552k regions, utilizing MethylSeq technology. The resulting sequencing data is processed through a standardized pipeline, which includes alignment and quality control (QC). The aligned BAM files are then subjected to methylation calling, read-wise methylation analysis, and fragmentomics. In the **ML Classification** stage, machine learning models are trained using a “leave-in” dataset and evaluated on a separate “leave-out” dataset. These models are designed to distinguish between cancer and control samples, encompassing a variety of cancer types, including colorectal, esophageal, gallbladder, liver, lung, pancreatic, stomach, breast, cervical, and ovarian cancers.

**Figure 3:**
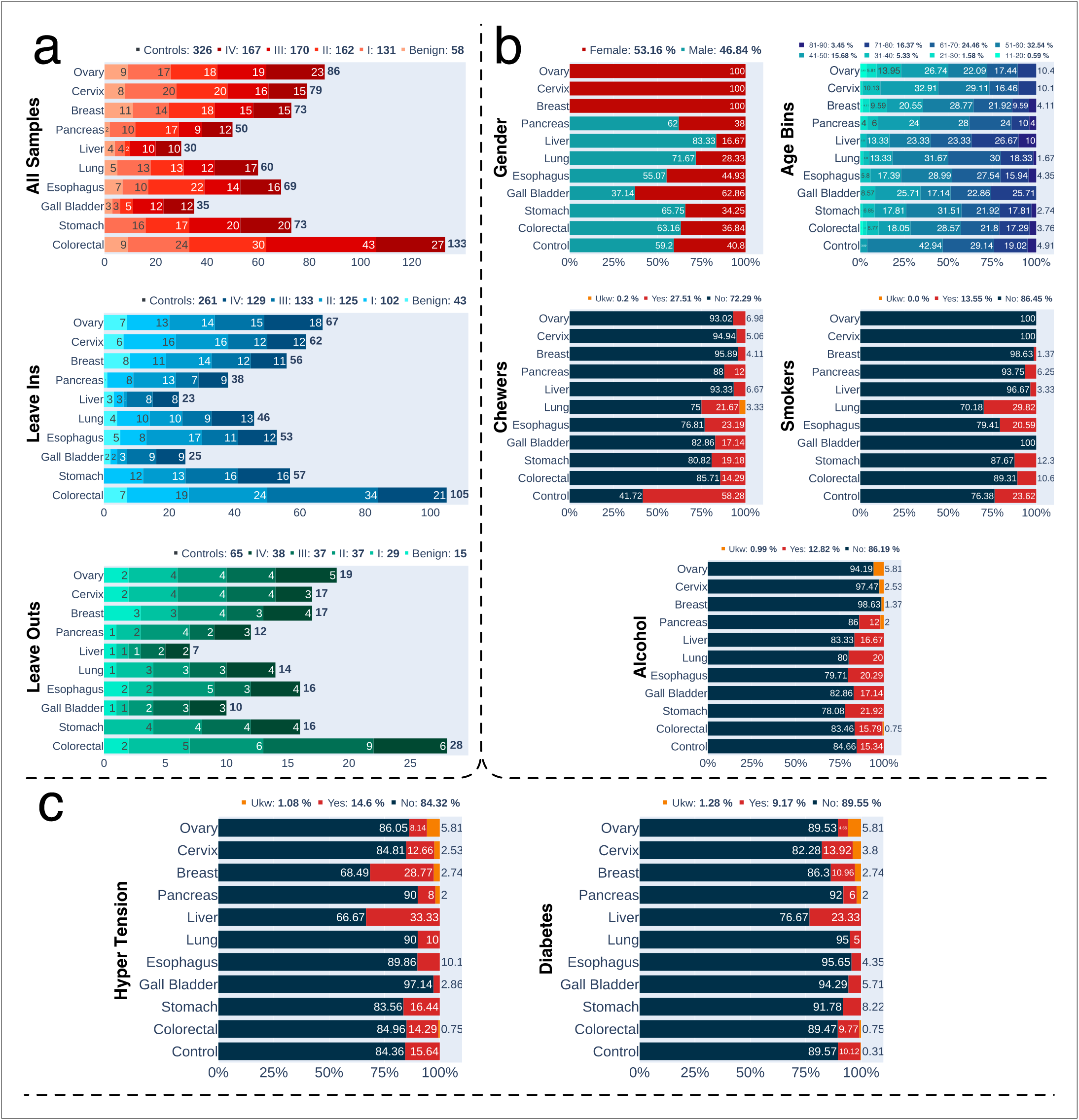
Cohort Characteristics and Distribution Across Cancer Types. Overview of key cohort characteristics **that include** age, gender, tobacco use, alcohol consumption, and comorbidities such as hypertension and diabetes. It is divided into four panels. **(a)** shows the distribution of **all** samples, **including** those in the “leave-in” and “leave- out” datasets, across various cancer types. Additionally, it presents a stage-wise distribution of samples within each cancer type. **(b)** shows the demographic distribution of gender, age bins, tobacco chewers, alcohol consumers, and smokers across various cancer types. **(c)** depicts the percentage distribution of lifestyle disorders like hypertension and diabetes across different cancer types.

### Signature of cfDNA specific metrics

Across 1,000 samples, an average of 215 million reads were sequenced per sample, achieving a mean coverage of 66x and on-target fraction 77% over the 123 Mb panel where the enzymatic conversion efficiency averaged 99%. After stringent filtering to retain on-target, uniquely mapped reads with low conversion errors, the final yield was approximately 120 million reads per sample.

We focused on four key features associated with circulating cell-free DNA (cfDNA) and cancer signatures, either derived directly from the assay or through computational analysis: a) Yield (ng/mL of plasma)[28] b) Fraction of fragments between 120-150 bp [29]c) Fraction of fragments with a 5’ end motif CCCA [30] , and d) Reads mapped to HPV16/18 [31]. Yield and the proportion of 120-150 bp fragments were expected to be higher in cancer patients compared to controls, particularly in later-stage cancers (Figure 4, Supplementary Figure 2). In contrast, the CCCA motif fraction was anticipated to be lower in cancer samples. For HPV, higher read counts aligned to HPV16/18 genomes were expected in cervical cancer samples.

**Figure 4:**
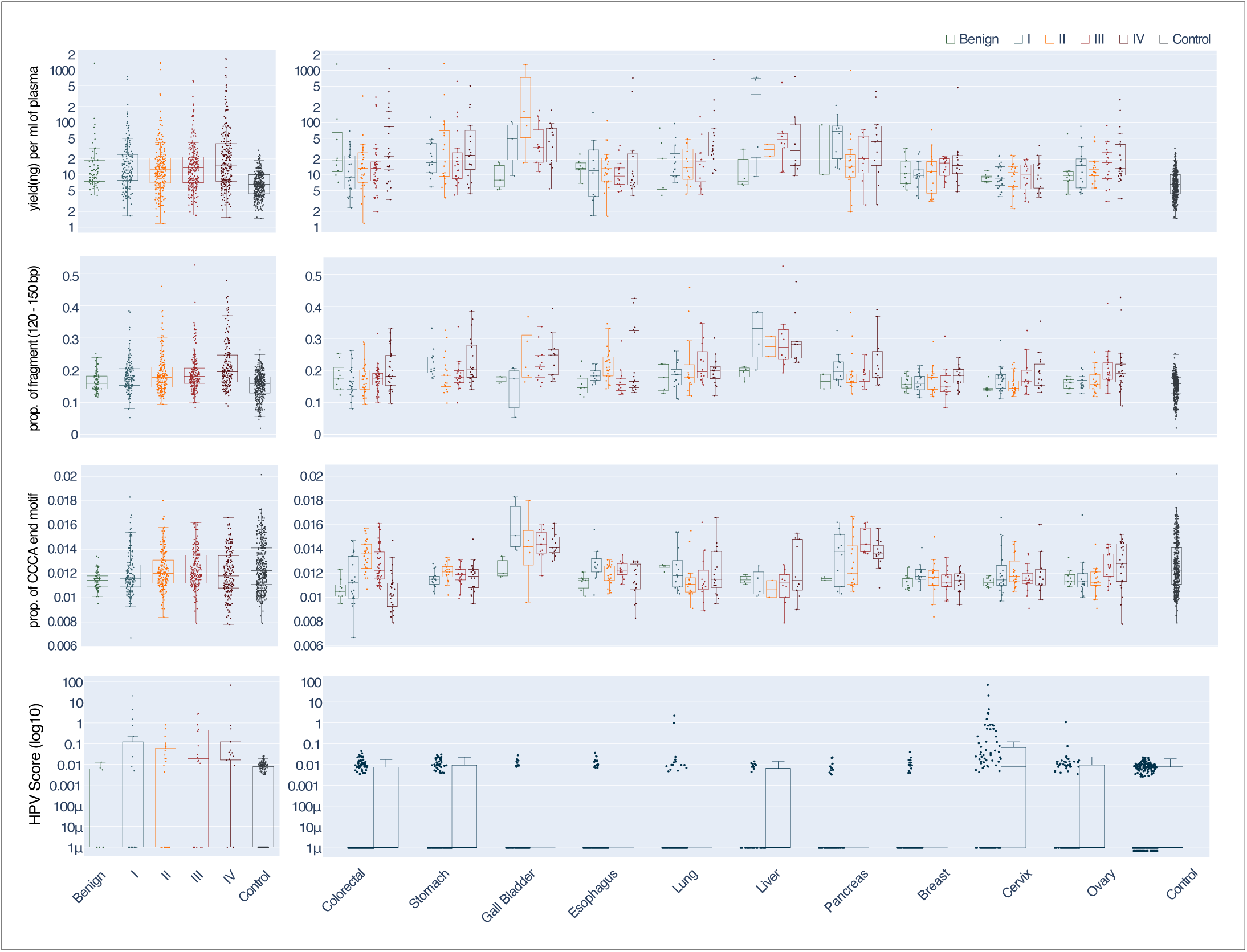
Distribution of Circulating Cell-Free DNA (cfDNA) Features Across Cancer Types and Stages. Key features associated with circulating cell-free DNA (cfDNA) and cancer signatures, derived either directly from the assay or through computational analysis: **Yield (ng/mL of plasma); Fraction of fragments between 120-150 bp; Fraction of fragments with a 5’ end motif CCCA; Reads mapped to HPV16/18**. For each feature, the variation is shown across different cancer types and stages, as well as in benign samples and controls. This analysis highlights the variability of cfDNA characteristics in relation to cancer and its potential utility in distinguishing between cancerous, benign, and control samples.

Cancer samples demonstrated significantly higher plasma yield and short fragment fractions compared to controls (t-test, *P<10e-7*). The enrichment of the CCCA motif was more prominent in controls, although this difference was less pronounced (t-test, *P=0.0003*). Notably, despite these overall trends, substantial variability was observed across different cancer types, suggesting that some cfDNA features may exhibit stronger signals in specific cohorts. A significant enrichment of HPV-mapped reads was detected in a subset of cervical cancer samples, with 47.25% of cervical cancer samples showing an HPV read fraction above 0.01 compared to 14.2% of samples above cut-off in the other cancer types and controls (fisher test *P<10e-7* , Figure 4).

### Performance of the cancer detection ML model

Detailed performance metrics for both the leave-in and leave-out sets are summarized in Supplementary Table 4, which also lists the number of regions selected for each of the two grouped models (see Materials and Methods). Briefly, the Control vs. Grouped Cancer model (7460 regions) demonstrated the following sensitivities in the leave-out set: 79.3% for Stage I cancers, 78.4% for Stage II, 78.4% for Stage III, and 86.8% for Stage IV, with an overall specificity of approximately 96.9% (Table 1). When optimizing outcomes, the gender-agnostic model was applied to Male samples, whereas both gender-agnostic and female-specific models were applied to Female samples. This approach resulted in an overall AUC of 0.96 (Figure 5a). Notably, lung and liver cancer samples achieved the highest sensitivity at 100%, while colorectal cancer samples showed a relatively lower performance at 54%. The entire ML workflow was repeated on random regions set (n=7460) to assess if they surpass the performance with de-novo regions. We observed that at specificity of ≥95% an overall sensitivity of 61% in Stage I and II (71% across all stages, Supplementary Table 7).

**Figure 5:**
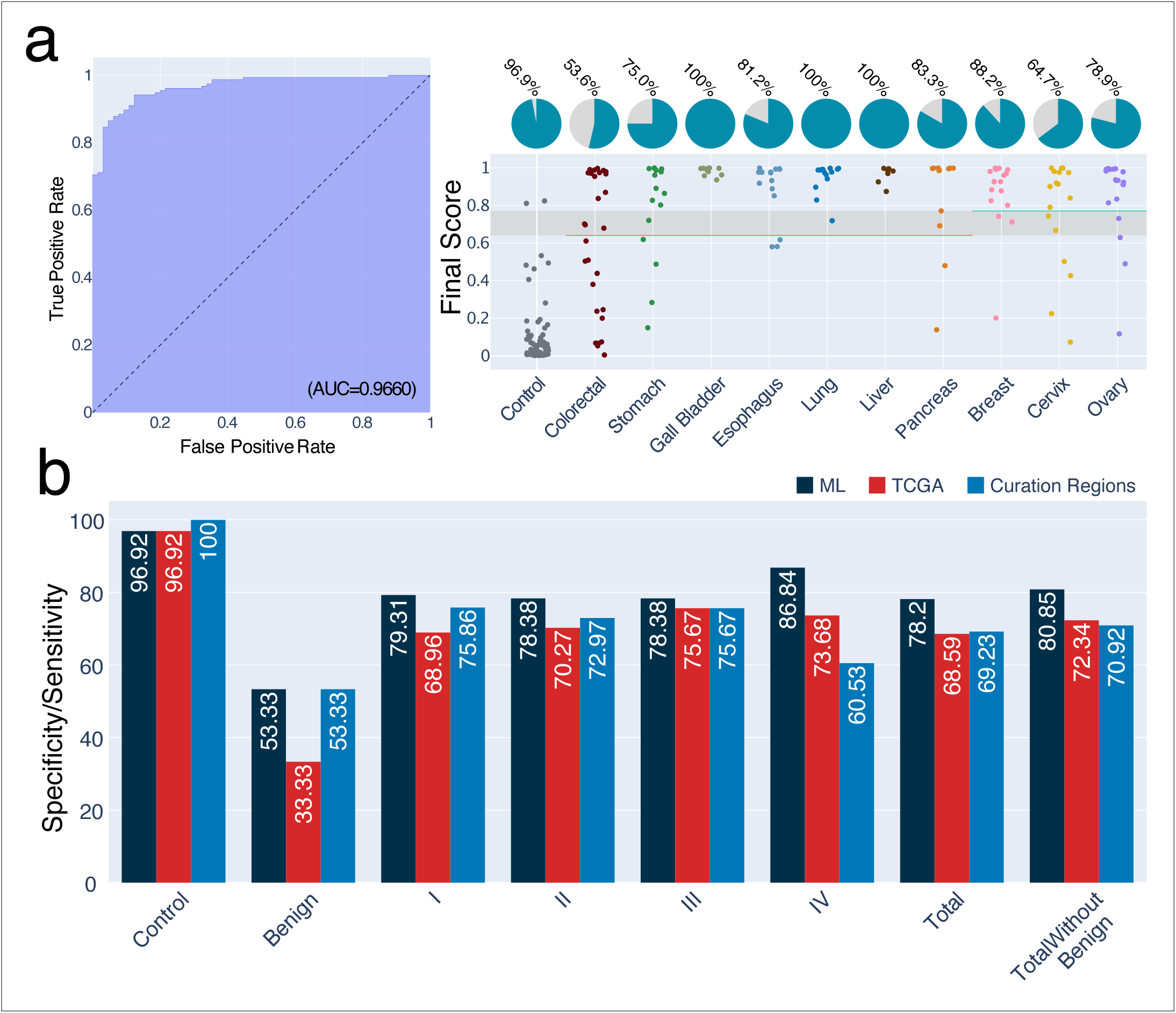
Model Performance. Overall model performance in classification of cancer and control samples **(a)** ROC curve for the grouped cancer detection model. The curve illustrates the model’s diagnostic performance, with an AUC of 0.966, indicating high accuracy in distinguishing between cancer and control samples. Additionally the distribution of final model scores and its performance across individual test samples is also Illustrated. The x-axis represents the control group and various cancer types, while the y-axis indicates the final score assigned by the model. Each dot corresponds to a test sample, with male samples evaluated using the General Cancer Model (colorectal, esophageal, gallbladder, liver, lung, pancreatic, and stomach cancers), and female samples assessed using both the General Cancer Model and the Female Cancer Model (breast, cervix, and ovarian cancers). The red and green horizontal lines mark the classification thresholds for the general and female cancer models, respectively. Samples below the threshold are classified as controls, while those above are classified as cancer. The pie charts depict the proportion of test samples correctly classified by the model, summarizing its overall performance. **(b)** Sensitivity and specificity across different cancer stages (I,II,III and IV), controls, and benign samples using three region sets: de novo ML-selected (7,460 regions), TCGA-derived (3,942 regions), and curated (2,459 regions).

**Table 1:** Cancer Detection (Control vs Cancer) performance on leave-out set.

| Ge80 + Yield (ng)/mL Plasma \| Control vs Group Cancer \| Evaluated at 95% leaveIn specificity |  |  |  |  |  |  |  |  |
| --- | --- | --- | --- | --- | --- | --- | --- | --- |
| LeaveOut specificity = 96.9% \| LeaveOut sensitivity = 80.8% |  |  |  |  |  |  |  |  |
| type | control | Benign | I | II | III | IV | Total | TotalWithoutBenign |
| control | 63 / 65 | 0 / 0 | 0 / 0 | 0 / 0 | 0 / 0 | 0 / 0 | 63 / 65 | 63 / 65 |
| Colorectal | 0 / 0 | 1 / 2 | 2 / 5 | 4 / 6 | 4 / 9 | 4 / 6 | 15 / 28 | 14 / 26 |
| Esophagus | 0 / 0 | 1 / 2 | 1 / 2 | 4 / 5 | 3 / 3 | 4 / 4 | 13 / 16 | 12 / 14 |
| Gall Bladder | 0 / 0 | 1 / 1 | 1 / 1 | 2 / 2 | 3 / 3 | 3 / 3 | 10 / 10 | 9 / 9 |
| Liver | 0 / 0 | 1 / 1 | 1 / 1 | 1 / 1 | 2 / 2 | 2 / 2 | 7 / 7 | 6 / 6 |
| Lung | 0 / 0 | 1 / 1 | 3 / 3 | 3 / 3 | 3 / 3 | 4 / 4 | 14 / 14 | 13 / 13 |
| Pancreas | 0 / 0 | 0 / 1 | 2 / 2 | 3 / 4 | 2 / 2 | 3 / 3 | 10 / 12 | 10 / 11 |
| Stomach | 0 / 0 | 0 / 0 | 4 / 4 | 2 / 4 | 4 / 4 | 2 / 4 | 12 / 16 | 12 / 16 |
| Breast | 0 / 0 | 2 / 3 | 3 / 3 | 4 / 4 | 2 / 3 | 4 / 4 | 15 / 17 | 13 / 14 |
| Cervix | 0 / 0 | 0 / 2 | 3 / 4 | 3 / 4 | 2 / 4 | 3 / 3 | 11 / 17 | 11 / 15 |
| Ovary | 0 / 0 | 1 / 2 | 3 / 4 | 3 / 4 | 4 / 4 | 4 / 5 | 15 / 19 | 14 / 17 |
| **Total** | **63 / 65** | **8 / 15** | **23 / 29** | **29 / 37** | **29 / 37** | **33 / 38** | **122 / 156** | **114 / 141** |
|  | **96.92%** | **53.33%** | **79.31%** | **78.38%** | **78.38%** | **86.84%** | **78.20%** | **80.85%** |

The performance of the Control vs. Cancer machine learning (ML) model was also assessed on samples with suboptimal enzymatic conversion efficiency (<99%). In these cases, downstream analysis was conducted using two approaches: one with the default set of high-quality aligned reads and another with an additional filtering step to exclude reads that exhibited low enzymatic conversion rates. Specifically, this filter targeted reads where non-CpG cytosines were not converted to thymine, as expected in mammalian genomes. The results indicate that incorporating this additional filtering (called as, “rescue”) step not only enhances the model’s tolerance for samples with low overall conversion efficiency but also improves performance. By focusing solely on highly converted reads, the model ensures that samples with poor conversion do not compromise classification accuracy. The model demonstrates a slight improvement in specificity, reducing the likelihood of misclassifying healthy samples ( Supplementary Table 8).

In parallel to the machine learning (ML) efforts to identify regions of interest, we conducted an extensive literature review and analyzed publicly available data to curate potential methylation markers (see Materials and Methods and Supplementary Document). Specifically, we leveraged differentially methylated regions (DMRs) from the TCGA dataset, focusing on hypermethylated regions within the top 1,000 ranked loci. This resulted in 3,231 DMRs, mapped to 4,782 regions within the Twist methylome panel. Applying the same ML pipeline and filtering using the Mann-Whitney U test, we narrowed this set to 3,942 regions. Additionally, a curated list of 3,765 regions derived from the literature and public databases was mapped to the Twist methylome and subsequently filtered during the ML step, yielding 2,459 regions. Performance comparisons of these regions— ML-selected (7,460 regions), TCGA-derived (3,942 regions), and curated (2,459 regions)—are shown in Figure 5b. At a specificity threshold of >95%, the ML regions outperformed both the TCGA and curated regions, with sensitivities of 81%, 72%, and 71%, respectively. Interestingly, there was limited overlap among the regions identified through the three approaches (Supplementary Figure 3). However, ML and TCGA-derived regions shared a significantly greater overlap compared to curated regions. Here, we took the intersection of the regions obtained from our analysis, TCGA hypermethylated regions, and curation regions (n=286) on which we observed a specificity of 95%, with the following sensitivities: Stage I: 58.6%, Stage II: 73%, Stage III: 81.1%, Stage IV: 73.7% (Supplementary Table 7).

### Tissue of origin model outcomes

The Tissue of Origin (TOO) models exhibited varying levels of sensitivity across their top-ranked predictions. The sensitivity for the Top I prediction was 57.9%, which increased to 77.2% for the Top II prediction and reached 80.7% for the Top III prediction. When analyzed stage-wise, the Top I model showed sensitivities of 52.2% for Stage I, 55.2% for Stage II, 72.4% for Stage III, and 51.5% for Stage IV. In contrast, the Top II model demonstrated higher sensitivity, with 78.3% for Stage I, 79.3% for Stage II, 82.8% for Stage III, and 69.7% for Stage IV (Table 2). Similarly, the Top III model achieved sensitivities of 78.3% for Stage I, 82.8% for Stage II, 86.2% for Stage III, and 75.8% for Stage IV. Further analysis presented in Figure 6, illustrates the accuracy of TOO predictions when considering the top two predictions for each sample, along with the patterns of misclassification. A prediction was deemed accurate if either of the top two predicted cancer types matched the actual tissue of origin. In cases where both predictions were incorrect, the top-ranked prediction was counted as a misclassification. Notably, the colorectal cancer cohort displayed the highest accuracy, while breast cancer samples showed the lowest rate of correct classification. These findings highlight differences in prediction performance across cancer types and suggest areas for further refinement, particularly in cohorts with lower accuracy (Table 2, Figure 6). Similar to the random analysis performed for the grouped model, random regions (count matched to the individual tissue of origin models) were used in the ML workflow for TOO which resulted in 67% accuracy for the Top II predictions. Genomic properties of the regions predicted to be important in both L1 and L2 are described in the Supplementary Document where we observe a majority of the regions (65%) fall in regions with the presence of enhancers which is significantly higher than that observed in the entire panel (46%).

**Figure 6:**
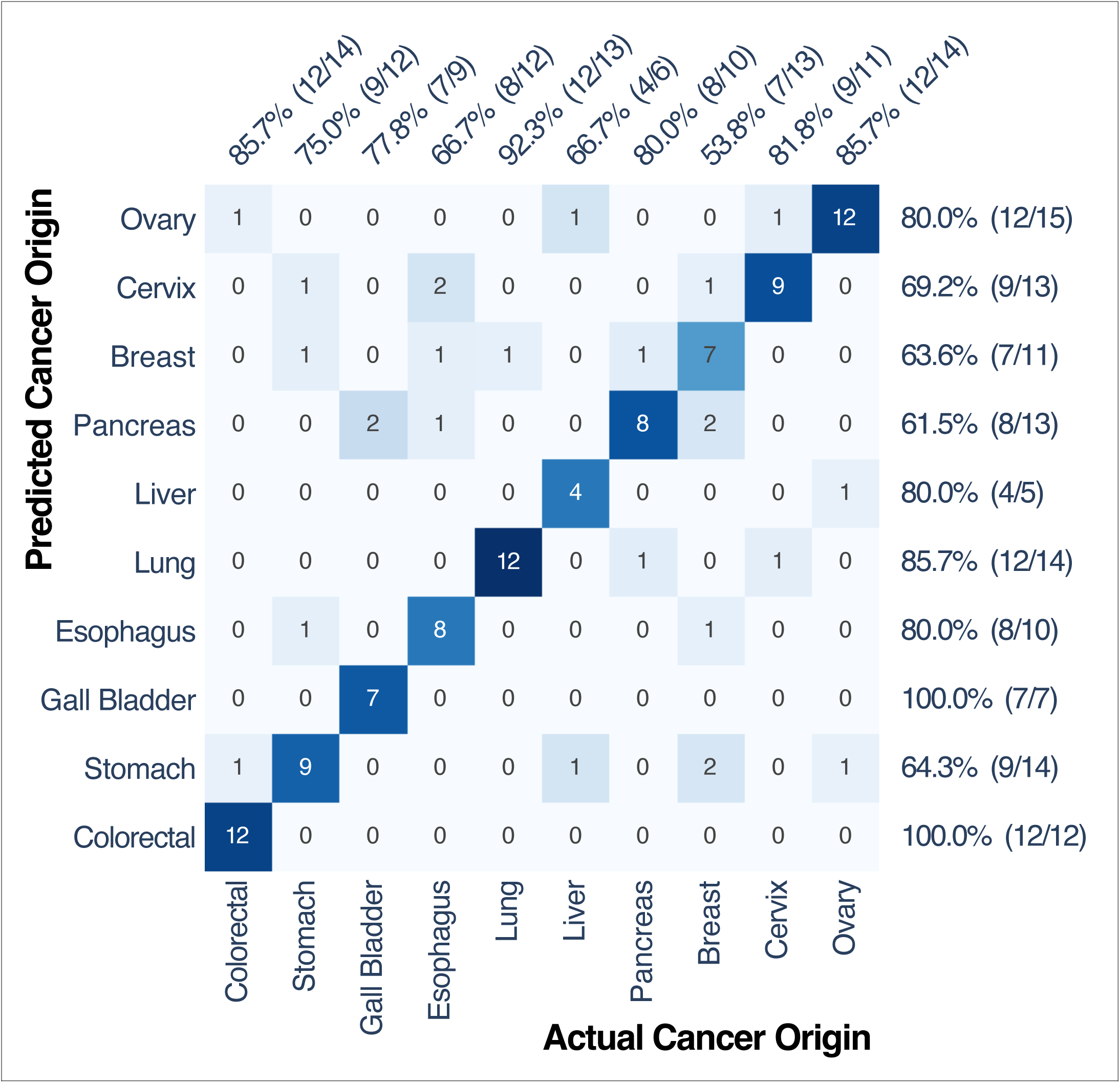
Accuracy of tissue-of-origin (TOO) predictions. Heatmap comparing the actual cancer origin (x-axis) with the predicted cancer origin (y-axis). Each cell represents the number of predictions, with color intensity indicating the level of concordance (diagonal cells) or discordance (off-diagonal cells). Diagonal cells highlight accurate predictions (if any of the top two prediction from model correctly identifies the tumor origin) while off-diagonal cells indicate mis-classifications (if the top two prediction from model doesn’t identify the tumor origin, the top most mis-classification is considered) showcasing instances where the predicted origin differed from the true origin. This visualization provides a comprehensive overview of the model’s performance in predicting tumor origins.

**Table 2:**
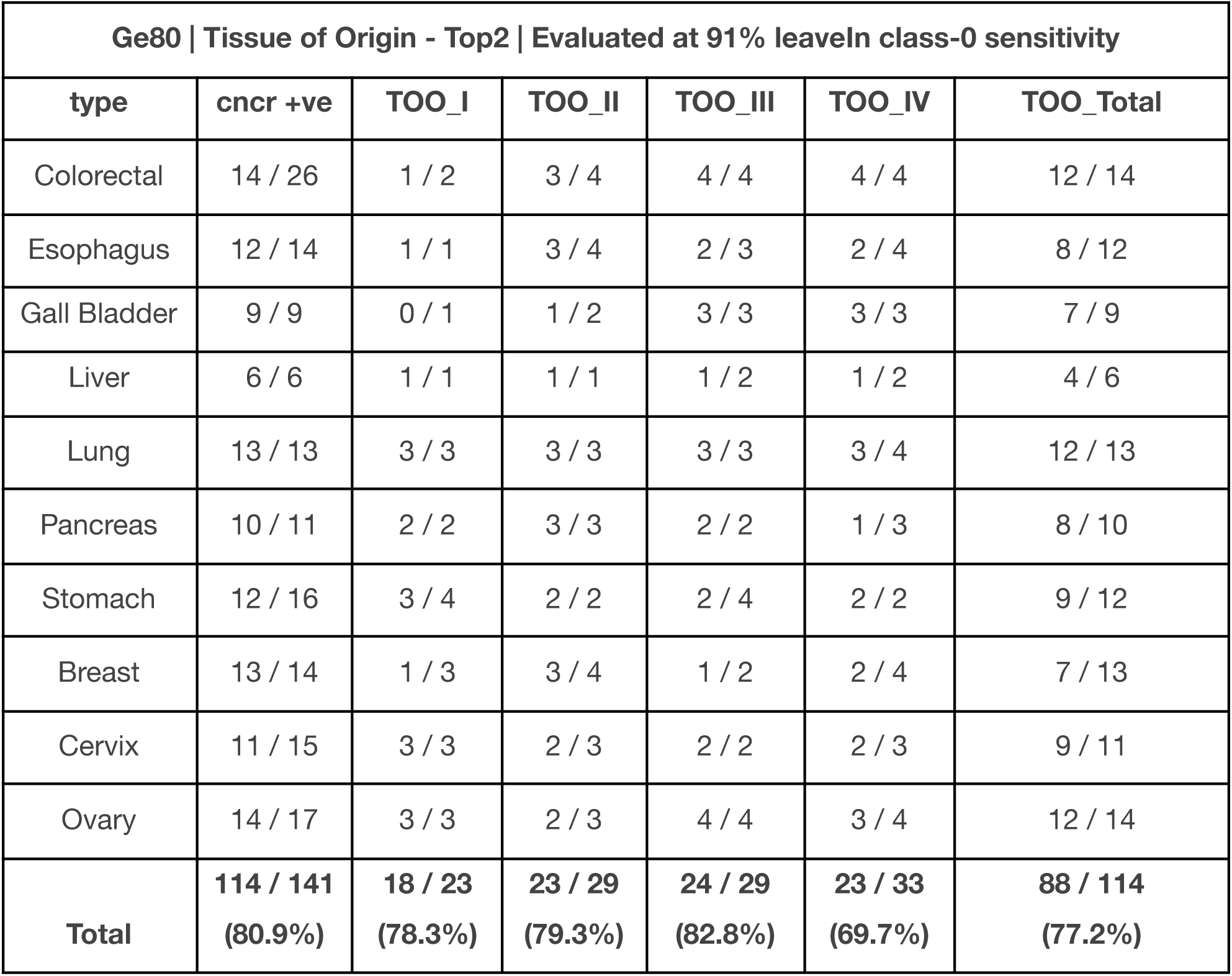
Tissue of Origin (TOO) Top2 performance on leave-out set.

| Ge80 \| Tissue of Origin - Top2 \| Evaluated at 91% leaveIn class-0 sensitivity |  |  |  |  |  |  |
| --- | --- | --- | --- | --- | --- | --- |
| type | cncr +ve | TOO_I | TOO_II | TOO_III | TOO_IV | TOO_Total |
| Colorectal | 14 / 26 | 1 / 2 | 3 / 4 | 4 / 4 | 4 / 4 | 12 / 14 |
| Esophagus | 12 / 14 | 1 / 1 | 3 / 4 | 2 / 3 | 2 / 4 | 8 / 12 |
| Gall Bladder | 9 / 9 | 0 / 1 | 1 / 2 | 3 / 3 | 3 / 3 | 7 / 9 |
| Liver | 6 / 6 | 1 / 1 | 1 / 1 | 1 / 2 | 1 / 2 | 4 / 6 |
| Lung | 13 / 13 | 3 / 3 | 3 / 3 | 3 / 3 | 3 / 4 | 12 / 13 |
| Pancreas | 10 / 11 | 2 / 2 | 3 / 3 | 2 / 2 | 1 / 3 | 8 / 10 |
| Stomach | 12 / 16 | 3 / 4 | 2 / 2 | 2 / 4 | 2 / 2 | 9 / 12 |
| Breast | 13 / 14 | 1 / 3 | 3 / 4 | 1 / 2 | 2 / 4 | 7 / 13 |
| Cervix | 11 / 15 | 3 / 3 | 2 / 3 | 2 / 2 | 2 / 3 | 9 / 11 |
| Ovary | 14 / 17 | 3 / 3 | 2 / 3 | 4 / 4 | 3 / 4 | 12 / 14 |
| **Total** | **114 / 141**<br>**(80.9%)** | **18 / 23**<br>**(78.3%)** | **23 / 29**<br>**(79.3%)** | **24 / 29**<br>**(82.8%)** | **23 / 33**<br>**(69.7%)** | **88 / 114**<br>**(77.2%)** |

### Validation of methylation signature

The de novo machine learning (ML) analysis for cancer vs. control and tissue-of-origin classification was compared against publicly available methylation signatures from The Cancer Genome Atlas (TCGA) across nine of the ten cancer types studied.Gallbladder cancer was excluded due to the lack of TCGA methylation data. We analyzed regions identified by both the L1-regularized grouped models and individual cancer vs. control models. While individual models were developed, the grouped model was preferred for final classification. For each identified region, we examined probe overlap with the Illumina 450K methylation array, the platform used by TCGA, and calculated the mean methylation level of all overlapping probes within the region.

The mean methylation level across all regions for a given model was used to determine the overall methylation status of each sample. Similarly, the average methylation across samples was calculated for both the cancer and the adjacent normal tissue cohorts to assess cohort-level methylation status. Our analysis consistently showed hypermethylation in cancer samples compared to controls within the selected regions from both L1 grouped models and standalone models for various cancer types. (Figure 7a, Supplementary Figure 4a, Supplementary Table 6). To further explore sample-level trends, we compared the distribution of methylation between cancer and control samples within TCGA cohorts. The results consistently demonstrated higher methylation levels in cancer samples compared to controls in the regions of interest (Figure 7b, Supplementary Figure 4b, Supplementary Table 6). It is worth noting that, in comparison, regions identified in the tissue-of-origin analysis did not reproduce distinct methylation signatures in the corresponding TCGA datasets for the respective cancer types (Supplementary Table 6).

**Figure 7:**
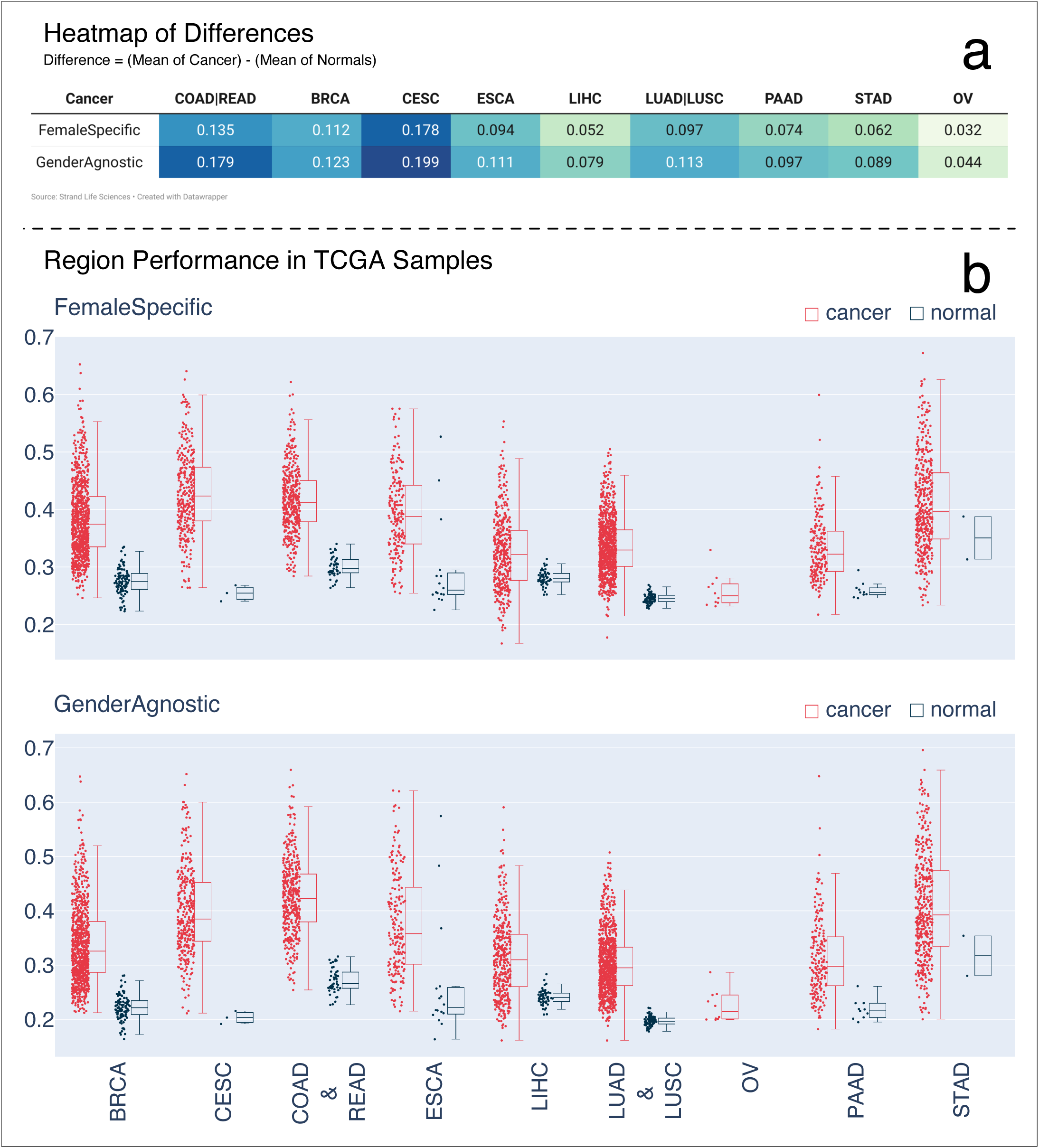
Methylation Distribution of Cancer and Adjacent Normal Tissues Across TCGA Cancer Types. **(a)** Heatmap where columns represent TCGA cancer samples, and rows correspond to regions showing differential methylation scores between each cancer type and its relevant control. Each cell reflects the difference in average methylation values between cancer tissue and adjacent normal tissue across all regions in the TCGA dataset. This visualization highlights the differential methylation patterns observed in cancer samples compared to adjacent normal tissues. (b) The x-axis represents different TCGA cancer types. For each cancer type, two plots are presented: one for the cancer tissue and one for the adjacent normal tissue. These plots display the distribution of mean methylation values for each group, highlighting the differences in methylation between cancer and adjacent normal tissues across the selected regions from the grouped models.

### Limit of detection and reproducibility

To assess the detection limits and robustness of our predictive models, we performed a systematic downsampling of PATR files, which record methylation status at the read level across the entire leave-out set. The objective was to determine how varying read depths impacted model performance, particularly in maintaining accurate predictions for both cancer detection and tissue-of-origin classification. During downsampling, we progressively reduced the number of reads and evaluated the accuracy of the models at each step. The results indicated that a minimum threshold of 100 million high-quality reads is critical for reliable predictions. High-quality reads were defined as those exhibiting efficient bisulfite conversion, specifically with a high proportion of non-CpG cytosines converted to thymine, indicating minimal technical artifacts. Maintaining this threshold was essential to preserve the integrity of the initial predictions derived from the complete dataset. Our findings showed that when the read count fell below 100 million, there was a noticeable decline in the model’s performance, particularly in identifying the tissue of origin. This suggests that tissue-specific methylation signals, which are inherently more variable, are especially sensitive to lower sequencing depths. In contrast, cancer vs. control classification remained relatively stable at slightly lower read counts but still benefited from maintaining the 100 million read threshold to ensure consistency and accuracy across all predictions (Supplementary Table 4). This analysis underscores the importance of sequencing depth and data quality in methylation-based models, highlighting that both sufficient coverage and accurate bisulfite conversion are prerequisites for dependable outcomes.

Finally, we evaluated the reproducibility of our pipeline by testing the impact of different sequencing platforms, multiple capture experiments, and repeated runs on the same platform using a given set of samples. The goal was to ensure that the assay and analysis pipeline consistently produced reliable results across varying conditions. Key metrics relevant to ML classification were assessed, including bisulfite conversion efficiency, on-target alignment rates, average global methylation levels, and region-specific methylated read fractions. These metrics were selected because they directly influence the performance of the classification models. Our analysis demonstrated that across all conditions—different platforms, repeated captures, and re-sequencing—the outcomes were highly comparable. Specifically, the observed consistency in methylation profiles and read alignment confirmed the robustness of both the assay and the pipeline in delivering reproducible and reliable results. This validation highlights the assay’s ability to maintain performance integrity despite technical variations, underscoring its suitability for clinical applications where reproducibility is critical (Supplementary Document).

## Discussion

Early detection of cancers offer advantages in survival rate and quality of life of individuals. Advances in NGS technology and enzymatic methylome sequencing have enabled seamless generation of methylation data with minimal DNA damage. cfDNA fragments originating from tumor cells bear methylation signatures of the primary cancer. The methylation patterns act as significant biomarkers in cancer detection. Since the cfDNA fragments of tumor origin are only a very small fraction of the cfDNA fragments in the sample, changes in average methylation patterns are very hard to detect. Instead, it is essential to measure and utilize the methylation patterns at the read/fragment level. Here we report the development and execution of an early detection test end to end; recruiting the subjects (controls and treatment-naive cancer samples), blood sample collection, plasma extraction, library preparation, sequencing, bioinformatics processing, read conversion assessment, methylation feature extraction, ML model building through region selection, and assessment of cancer status with the likely tissue(s) of origin.

Given the inherently low tumor fraction in circulating cell-free DNA (cfDNA), we emphasized the critical need to identify well-converted reads and fragments to ensure accurate downstream analysis. To address this, we developed a customized algorithm that specifically evaluates the conversion efficiency of non-CpG cytosines. By selectively retaining only high-quality, fully converted reads, this approach effectively minimized the risk of false positives due to incomplete bisulfite conversion, which can artificially inflate methylation signals and obscure true biological patterns. Recognizing the importance of robust coverage in critical regions, we focused on regions with consistent, high-depth reads. We also implemented synthetic data augmentation and region-specific selection to enhance the robustness of our machine learning (ML) models. This strategy was essential for mitigating feature variability caused by technical differences, such as sequencing platform changes and batch effects, ensuring that the models could generalize across diverse conditions. Our ML pipeline operates at two hierarchical levels. The first level targets cancer detection, achieving high specificity and sensitivity in identifying samples with cancer-associated methylation profiles. The second level refines the analysis to predict the tissue(s) of origin. To further reduce false positives, we incorporated gender-specific models, which allowed for a more precise subset of models to be evaluated based on the sample’s characteristics.

This comprehensive, hierarchical approach resulted in a balanced performance profile, with models achieving over 78% sensitivity for early-stage cancer detection and a specificity exceeding 97%. For positive samples, tissue-of-origin (TOO) classification reached 77% accuracy with a specificity of 91%. These results underscore the pipeline’s efficacy in delivering both accurate cancer detection and reliable identification of the tissue of origin, effectively tailored to the dataset’s characteristics and underlying cancer biology. We further demonstrated the superior sensitivity of our models, both for cancer vs. control and TOO predictions, compared to count-matched random sets derived from the same methylation panel. Notably, the sensitivity for early-stage cancer detection improved by approximately 17% when using our selected regions of interest (ROIs) over random regions, highlighting their predictive value. However, it is important to note that the Twist methylation panel we employed was pre-designed to capture pan-genome methylation hotspots. As a result, our initial set of ∼552K regions was already biased towards regulatory regions implicated in tumorigenesis, which explains why even random sets yielded a baseline sensitivity above 50%. In addition to assay development for early cancer detection—marking a pioneering effort for an Indian patient cohort—we conducted a rigorous analysis to identify differentially methylated regions (DMRs) from publicly available datasets, such as TCGA, and performed extensive literature curation to compile a list of cancer-associated markers. We evaluated the ML performance of these curated regions alongside regions identified through our in-house analyses. Our findings revealed that effective cancer-control separation can emerge from diverse strategies, including public methylation data, community-driven marker curation, and novel patient-control comparisons. To assess the combined utility of these strategies, we created an intersected list of regions from in-house datasets, TCGA-derived regions, and curated markers, then re-ran the ML workflow. Models built solely on de novo regions consistently outperformed those using orthogonal marker sets by a small but significant amount ( a 4–10%). This highlights the value of integrating orthogonal methods for marker selection. A targeted panel incorporating the most informative features from each strategy could further enhance overall detection performance, potentially optimizing sensitivity and specificity across a broader spectrum of cancer types.

Finally, we evaluated the methylation status of our machine learning-predicted regions of interest (ROIs) from cfDNA derived from blood plasma against tumor biopsy data from TCGA. We observed a consistent trend of hypermethylation in these regions across TCGA tumor samples compared to adjacent normal tissues. This finding reinforces the biological relevance of our selected regions in distinguishing cancerous from non-cancerous samples. However, the regions identified as significant for tissue-of-origin (TOO) classification did not show a similar consistent methylation pattern in TCGA, indicating that TOO-specific markers may not overlap directly with hypermethylation patterns observed in public datasets. One could postulate here that methylation alterations in cancer genomes are widespread and not necessarily confined to discrete, tumor-specific regions. Instead, it appears that tumor-specific signatures may be less pronounced and are likely characterized by subtle, combinatorial patterns rather than isolated methylation events. The ML algorithms effectively leverage these unique combinations of pervasive methylation changes to accurately predict the tissue of origin, highlighting the complex, multi-region nature of methylation landscapes in cancer. This underscores the utility of ML in identifying nuanced patterns that may not be evident through traditional single-region analyses, emphasizing its role in enhancing tissue-specific classification based on global methylation profiles. In summary, we present a comprehensive end-to-end workflow, encompassing sample acquisition, cfDNA extraction, and computational classification, capable of detecting cancers at early stages. This study highlights the clinical utility of our approach, marking a significant milestone by demonstrating its effectiveness in an Indian population cohort for the first time. Our findings underscore the potential of such a product in enhancing early cancer detection and guiding clinical decision-making in diverse healthcare settings.

## Authors’ contributions

**S. Basu:** Investigation, supervision, data analysis, QC, manuscript-review and editing. **P. Hiremath:** Data Analysis, pipeline development, fragmentomics, ML modules, manuscript–original draft, manuscript-review and editing. **N. Rathod:** Data Analysis, signal purification modules, ML modules, manuscript-figures and editing. **A. Chatterjee:** Ethics documentation and queries, sample acquisition, QC-metadata. **D. Vishwanath:** Laboratory workflow, protocol standardisation. **A. Ghosh:** Sample tracking, record digitisation, metadata-followup. **S. Sanguri:** Data review, demographics, habits analysis. **S. Chakraborty:** QC, pipeline development, pipeline execution. **A. Tripathi:** QC, standardization, reproducibility, spot-checks. **RT. Preetha, A. Nair, G. Kumar:** Plasma/cfDNA extraction, library preparation, capture. **K. Sekar:** Cloud Infrastructure, pipeline execution, QC-dashboards. **S. Yete, G. Bhanumathy, U. Bahadur:** Literature survey of methylation markers. **A. Radhakrishnan:** Case Report form, consent form, hospital onboarding, open clinica designing. **A. Khan:** Library preparation and capture. **Y. Kannan S:** QC-targeted panel, ML signals in TCGA. **L. Bollipalli:** LIMS-clinical metadata. **P. Ghana:** Sample collection, project management. **A. Ramanathan, P. Saha:** Project management. **S. Phalke:** Lab protocol development. **C. Cantor:** Scientific advisor. **S. Limaye:** Consulting Oncologist. **V. Chandru:** Scientific advisor. **V. Veeramachaneni, R. Hariharan:** Investigation, lab metrics, ML advisor, project design.

## Data Availability

All data produced in the present work are contained in the manuscript and it's supplementary figures/tables.

## Acknowledgments

**L. Alagappan, S. Parida, R. Sathasivam, C. Totagi, V. Kumar NS, A. Bhandari, N. Lakshmaiah, N. Kagal, M. Prakash, P. Manoja:** The authors would like to express their gratitude to these individuals whose support and contributions were essential to the successful completion of this study.

The authors sincerely thank the principal investigators and the patients who generously participated in this study under Clinical Trial Registration No: CTRI2022/05/042936. Their contributions and commitment were invaluable to this study.

## Supplementary Data

Suppmentary Document Description of methods for sample collection, lab and analysis protocols, read conversion assessment, sample reproducibility, region characteristics, public methylation data analysis.

Supplementary Table 1 This table summarizes the distribution of control and cancer samples across different sites, along with staging information for the following cancer types: breast, esophageal, ovarian, lung, stomach, colorectal, cervical, and hepatobiliary.

Supplementary Table 2 This table offers a comprehensive summary of the sample cohort, including stage-wise distribution for each cancer type. It also presents demographic and clinical statistics, such as gender distribution, mean age, BMI, and lifestyle factors (e.g., chewing, smoking, alcohol consumption). Additionally, it reports the prevalence of lifestyle disorders, including hypertension and diabetes, across various cancer types. The table further includes sample counts from different anatomical sites and metadata for a total of 1,014 samples.

Supplementary Table 3 This table provides the distribution of different cancer types, classified by stage (I, II, III, IV) and benign status in both the leave-in and leave-out datasets. It also includes cancer and stage-specific percentage statistics for females, chewers, and mean age across both datasets.

Supplementary Table 4 This table presents the performance statistics of the machine learning models, including key metrics such as sensitivity and specificity.

Supplementary Table 5 This table provides information on curation regions, TCGA seed probes, and TCGA differentially methylated regions (DMRs).

Supplementary Table 6 This table provides a comprehensive summary of methylation statistics in TCGA cancer samples for the regions obtained from our analysis.

Supplementary Table 7 This table provides the ML performance for the models built using TCGA regions, curation regions, intersection of regions (Strand de-novo, TCGA, and curation regions), and random regions.

Supplementary Table 8 This table presents the performance metrics of the machine learning model, comparing results before and after the rescue of good converted reads, where non-CpG cytosines (C’s) are converted to thymine (T’s).

## Supplementary Document

### 1. Sample collection from centre

#### Subject Enrolment

Treatment naive patients with confirmed (i) malignant tumors including but not limited to breast, cervix, colorectal, esophagus, lung, oral, stomach, ovarian cancers who were eligible for either surgical tumor resection and/or treatment for carcinoma (ii) patients with benign and precancerous neoplasm/lesions for the anatomical sites mentioned mentioned above. Clinical diagnosis by the treating oncologist based on their routine clinical practice and SOC is accepted.

. Prior to initiation of recruitment, ethical approval was obtained from each site for the study for that site. Each patient were approached by the site Principal Investigator or designees to obtain consent for study participation as detailed below:

● A description of the objectives of the study and how it was organized
● The type of tests and procedures in the study
● Any potential negative effects attributable to the study
● The freedom to ask for further information at any time
● The subject’s right to withdraw from the study at any time without giving reasons and
● without jeopardizing the subject’s further course of medical treatment
● The existence of payment for medical care of the subject
● Adequate time and opportunity to get questions answered

Subjects had the opportunity to seek external consultations and had any questions answered before signing the informed consent form (ICF). Each ICF was also signed by the investigator or designee (with date). If the subject was unable to read/write, an impartial witness signed (with date) the ICF to attest that the details were accurately explained to and apparently understood by the subject. A copy of the signed ICF was shared with the subject. The following bio-specimens were collected from the subjects after receiving their informed consent. Blood from subjects with confirmed malignant tumors , benign and precancerous cases/lesions was collected only from patients who had not undergone any prior treatment regimens (treatment naïve) from an arterial line and/or venous line .

The following informations were collected and recorded in the case report form (<u>CRF</u>):

1. Demographics (Age, Gender, Date of birth, Contact details)
2. Baseline physical (height and weight)
3. Vital signs

● BP (systolic and diastolic, mm Hg),
● Heart rate (beats per minute),
● Body temperature, and
● Respiratory rate
4. Clinical history:

● Disease history
● Treatment history (if any)
● Allergy and nutrition history
5. Current diagnoses, including current treatment, medications, interventional procedures and comorbid conditions, if any.
6. Data pertaining to the family history, or menarche age, childbirth and lactating period for gynecological cancer
7. Redacted pathological and radiological reports

#### Sample Transportation

Collected blood in Streck tubes were transported at room temperature within 48-72 hrs of sample collection from the site and tracked through a Sample tracking log.

### 2. Alu-PCR estimation of methylSeq conversion

Quantification and quality assessment of fragmented DNA (FFPE DNA and cfDNA) can be done by a quantitative PCR (qPCR) method to amplify the Alu elements in the human genome (<u>Ref</u>). The Alu sequences are ubiquitous in the human genome and thus be used as a universal control across all human samples. Similarly, we can also use the Alu qPCR assay to determine the percentage of conversion of unmethylated cytosines to thymines in a MethylSeq library. Unfortunately, there is no standard method to assess the quality of conversion prior to sequencing the MethylSeq libraries, making the Alu qPCR approach a valuable tool in this context. Serial dilutions of human genomic DNA (Cat No.:G304A, Promega Corporation) were made with nuclease-free water to yield concentrations of 3.3, 0.33, 0.033, 0.0033, 0.00033, and 0.000033 ng/μl, which constituted the standard curve. Unknown samples were also diluted to an approximate concentration of 10pg/μl. The qPCR reaction mix was then prepared according to the following table:

| Component | Volume for<br>1 reaction<br>(μl) |
| --- | --- |
| SYBR Fast qPCR master mix (2X)<br>(Kapa) | 10 |
| Alu 115 forward primer 10μM | 1 |
| Alu 115 reverse primer 10μM | 1 |
| Nuclease-free water | 4.6 |
| Low RoX | 0.4 |
| Total | 17 |

For the standard curve, as well as the unknown samples, 17μl of the above master mix and 3ul of the DNA dilution was added. Appropriate positive and negative controls were included in the assay. The standards and samples were loaded in strip tubes or 96-well plates, which were then placed in the qPCR system (AriaMx, Agilent technologies); the qPCR program mentioned below was run:

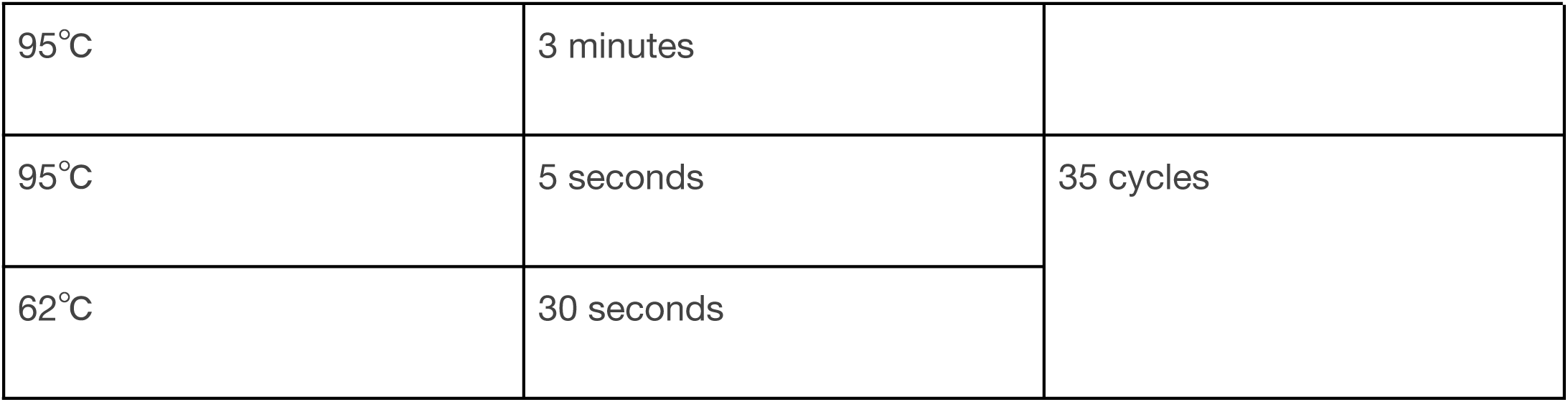

The Ct values obtained for each well for the standards and the unknown samples are then tabulated against each sample name. An average of the values obtained from the three wells is calculated to obtain the average Ct value. It can be seen that samples that have a Ct value equal to or greater than 21, then the MethylSeq libraries are seen to have a % conversion (Non-CpG) and % conversion (Lambda DNA) greater than 99%. Samples with Ct values lesser than 21 are seen to have poor conversion post sequencing. A standard curve was plotted with the Ct values obtained on the y-axis versus the concentration values on the x-axis in the log scale, as shown below.

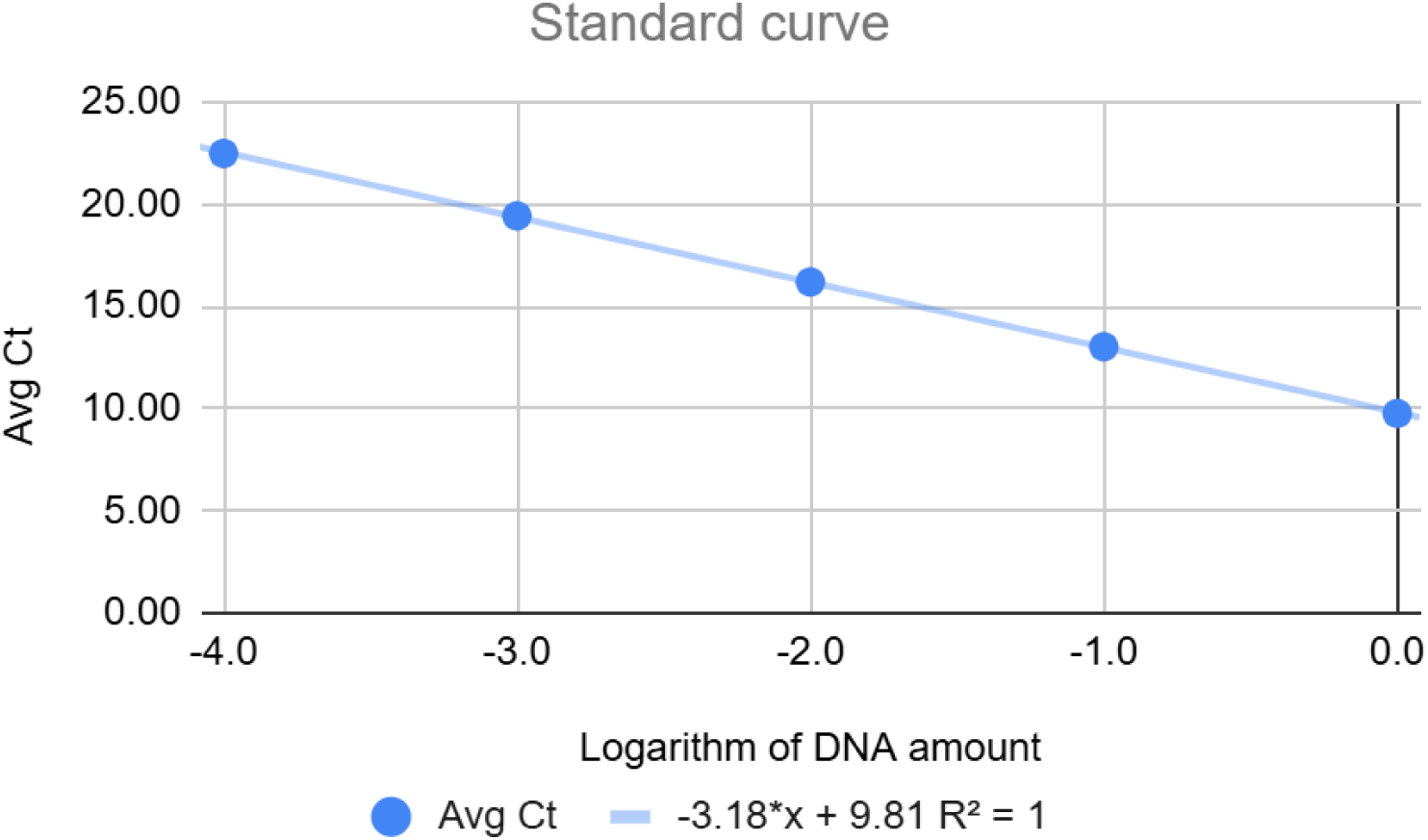

The slope and the y-intercept are identified. The Ct values for the unknown/test samples are then extrapolated using the below equation to get the amount of input DNA actually detected in the sample wells.

Amount in ng = ( 10^(Ct – y-intercept) ) / slope

% amplifiable DNA can be calculated using the below formula:

(Amount in ng from extrapolation in standard curve / Amount in ng of the sample actually added)*100

If % amplifiable is <0.5, then the library is seen to have good % conversion

### 3. Conversion of unmethylated cytosines at a read level

1. Take a double stranded fragment f that is sequenced.

2. f yields two reads, say r1 mapping to the reference and r2 mapping to the rev comp of the reference.

3. Trace f’s PCR lineage back to an original double stranded molecule X.

4. Let Xs be the strand of X that yields f.

5. Then f carries conversion info for cytosines in Xs but not in Xs’. (Xs’ is the other strand)

6. Two cases now, assuming Xs is either fully converted or not

a. If Xs is in the same direction as the reference

I. If the nonCpG Cs in the reference align with Cs in r1 and likewise non CpG Cs in the reference align with Gs in r2, then f is not converted

1. ii. If the nonCpG Cs in the reference align with Ts in r1 and likewise non CpG Cs in the reference align with As in r2, then f is converted

b. if Xs is in the opp direction to the reference

i. If the nonCpG Gs in the reference align with Gs in r1 and likewise non CpG Gs in the reference align with Cs in r2, then f is not converted

ii. If the nonCpG Gs in the reference align with As in r1 and likewise non CpG Gs in the reference align with Ts in r2, then f is converted

7. How do we know which of the above two cases applies for Xs?

a. We don’t, so we try each of the two cases

b. If Xs is fully converted, then f will satisfy either 6aii or 6bii but not both

c. If Xs is not converted at all, f will satisfy both 6ai and 6bi

d. For 6a, we need the reference to overlap enough non CpG Cs in r1 U r2 (say >=3)

e. For 6b, we need the reference to overlap enough non CpG Gs in r1 U r2 (say >=3)

f. We drop fragments that do not satisfy both the above, and then those that do not satisfy one of 6aii or 6bii (up to 1 exceptions)

g. Claim:

i. The fraction of fragments f that remain is ∼ conversion efficiency calculated locus wise from the CHH/CHG bedgraph file

i. The distribution of such fragments along the genome is not biased relative to the distribution of all fragments

### 4. Sample reproducibility

The reproducibility of sequencing across different runs and platforms was evaluated using three combinations of sample conditions.

The first combination involved samples from the same subject ID, time point, extraction time point, library preparation time point, and capture pool time point, which were sequenced on two different occasions using different sequencers. Specifically, the samples were run on the NovaSeq 6000 and NovaSeq X Plus platforms to assess consistency in sequencing performance. Various sequencing quality parameters were compared, including the number of reads sequenced (in millions), percentage of duplicate reads, percentage conversion of non-CpG cytosines, on-target alignment rates, and the percentage of methylation calls. These metrics provided insights into the reproducibility of sequencing results across platforms, ensuring that the observed outcomes were not significantly influenced by the sequencing system used.

In Figure 4.1, we compare the sequencing results across different runs, where Run0303 represents sequencing on the NovaSeq 6000 platform, while the remaining three runs were performed on the NovaSeq X Plus platform. This comparison highlights the performance differences between the two sequencing platforms across several key metrics. The Ge80 Pearson correlation was calculated between Run0303 and Run0061 since they show the significant difference when the same sample was run on different sequencers.

In Combination 2, samples from the same subject ID, time point, extraction, library preparation time point, and capture pool time point were sequenced on two separate occasions using the NovaSeq X Plus platform. This comparison aimed to evaluate run-to-run consistency on the same instrument. Key sequencing metrics, including the number of reads, percentage of duplicates, conversion of non-CpG cytosines, on-target alignment, and methylation percentage, were analyzed to assess reproducibility.

Figure 4.2a highlights the variations in these sequencing metrics when the same set of samples was sequenced across two separate runs on the NovaSeq X Plus. The metrics compared—percentage of duplicates, non-CpG cytosine conversion rates, on-target alignment, and methylation percentage—provide insights into the degree of variability observed within the same sequencer.

Figure 4.2b depicts the variability in key sequencing metrics—number of reads, percentage of duplicates, percentage conversion of non-CpG cytosines, on-target alignment (pAlnOT), and average coverage (cov.avg)—when the same set of samples was sequenced across different runs. Among these runs, Run0061 and Run0067 were performed using the 25B flow cell, while Run0064 utilized the 10B flow cell. This comparison also provides insight into how flow cell type influences sequencing performance and consistency.

In the third comparison, samples originating from the same subject ID, time point, extraction time point, and library preparation time point were divided into two different capture pools, processed at separate time points. This setup aimed to assess the impact of capture pool variability on sequencing outcomes. By comparing these samples, the analysis focused on determining the extent to which differences in capture pool preparation contribute to variations in key sequencing metrics.

Figure 4.3a illustrates the sequencing metrics for samples processed across various capture pools. Notably, the samples in Poolib-82-TRGS_Poolib-101-TRGS_Poolib-100-TRGS are 8 plex samples whereas in PoolLib-146-TRGS and PoolLib-147-TRGS are 16 plex and 24 plex respectively. This variation in plex levels provides additional context to assess how capture pool configurations influence sequencing performance.

Figure 4.3b indicates the differences in the number of reads obtained from sequencing, percentage duplicates, conversion efficiency, on target alignments, average coverage and percentage methylation when the same sample was sequenced in different pools.

**Figure 4.1:**
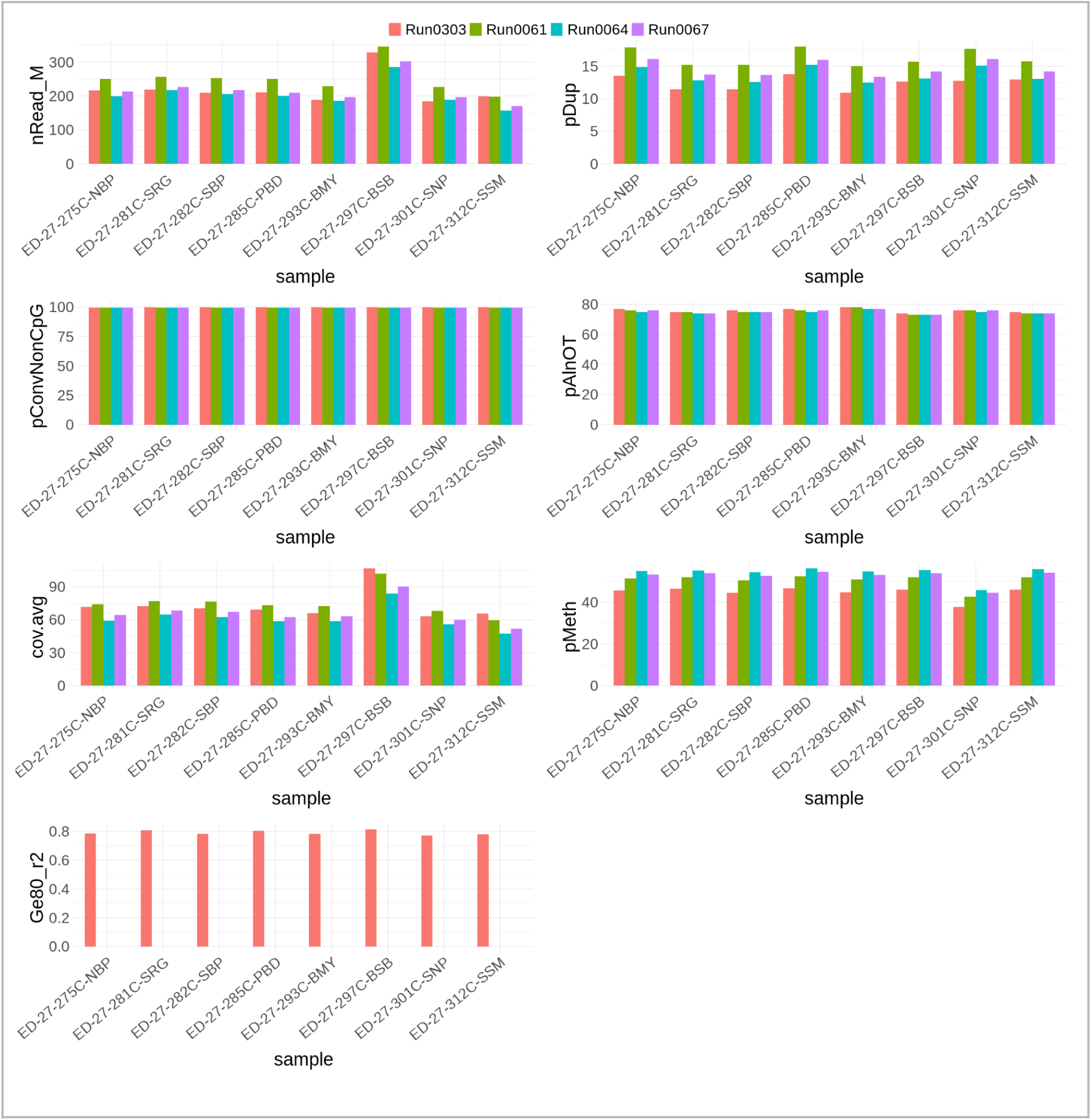
Reproducibility in case of different sequencer i.e NovaSeq 6000 and NovaSeq X Plus sequencer on the same set of samples

**Figure 4.2a:**
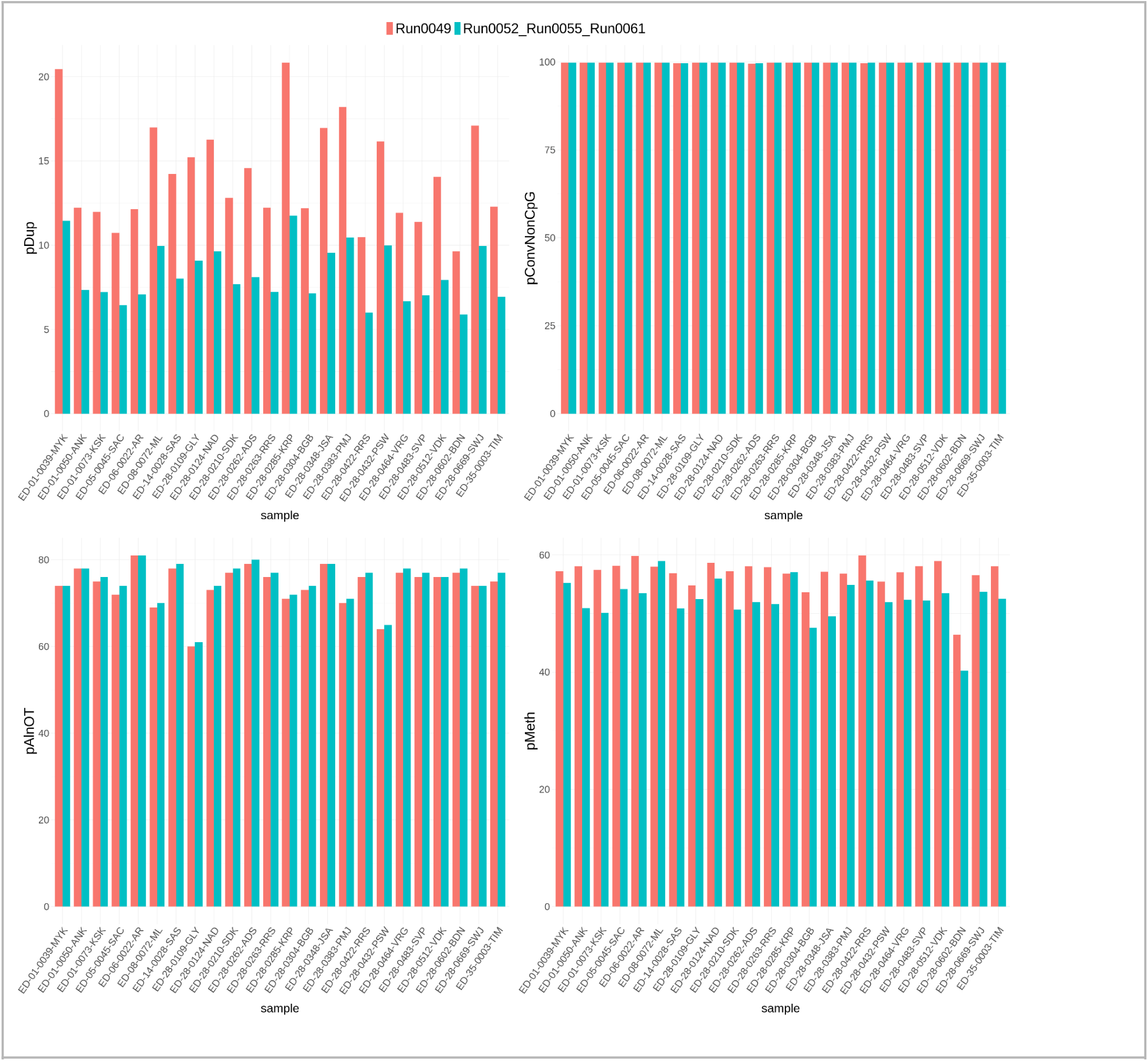
Reproducibility when the same set of samples are run on different occasions.

**Figure 4.2b:**
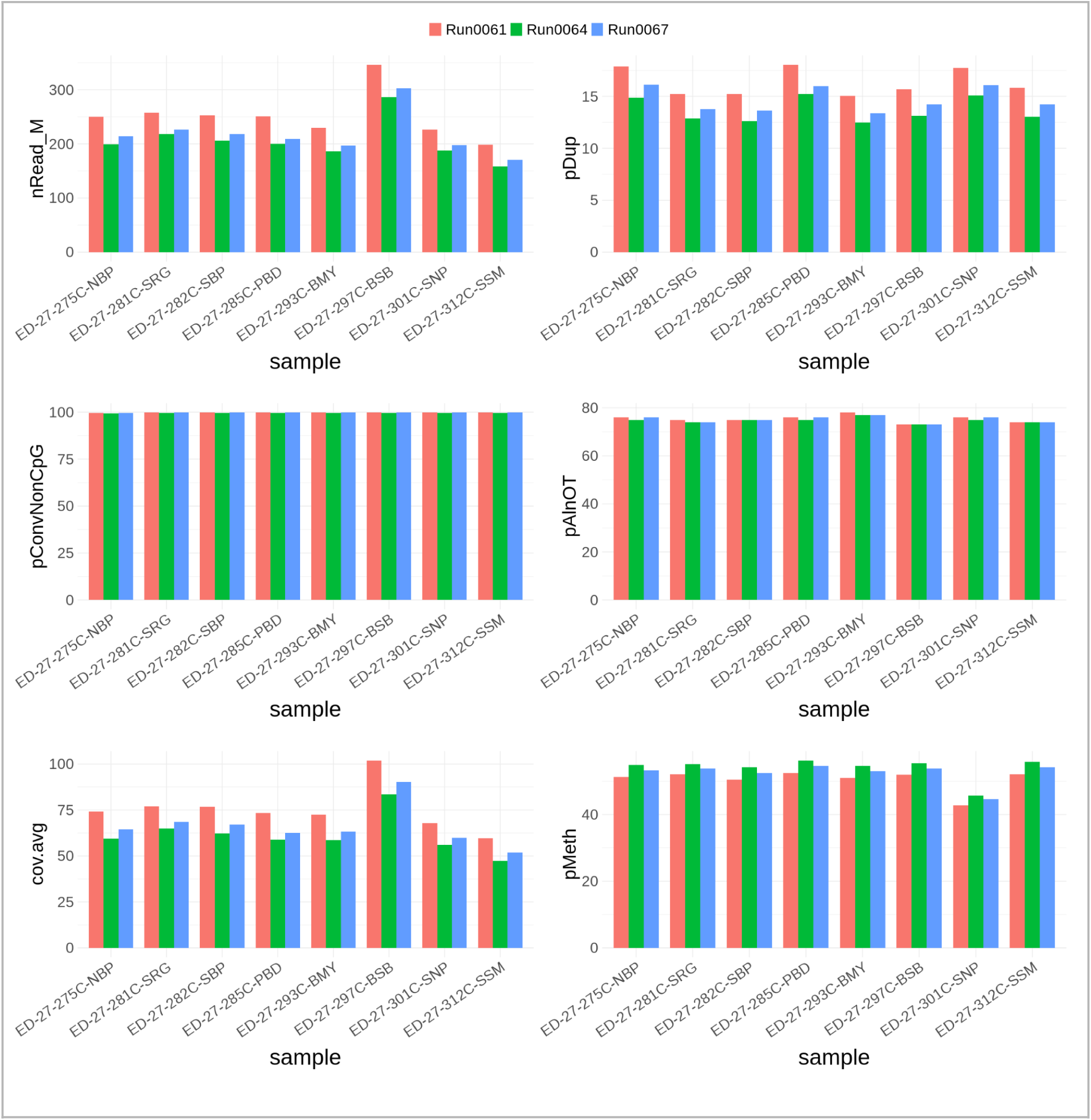
Reproducibility when the same set of samples are run on different occasions, with some variability being introduced by the difference in flow cell.

**Figure 4.3a:**
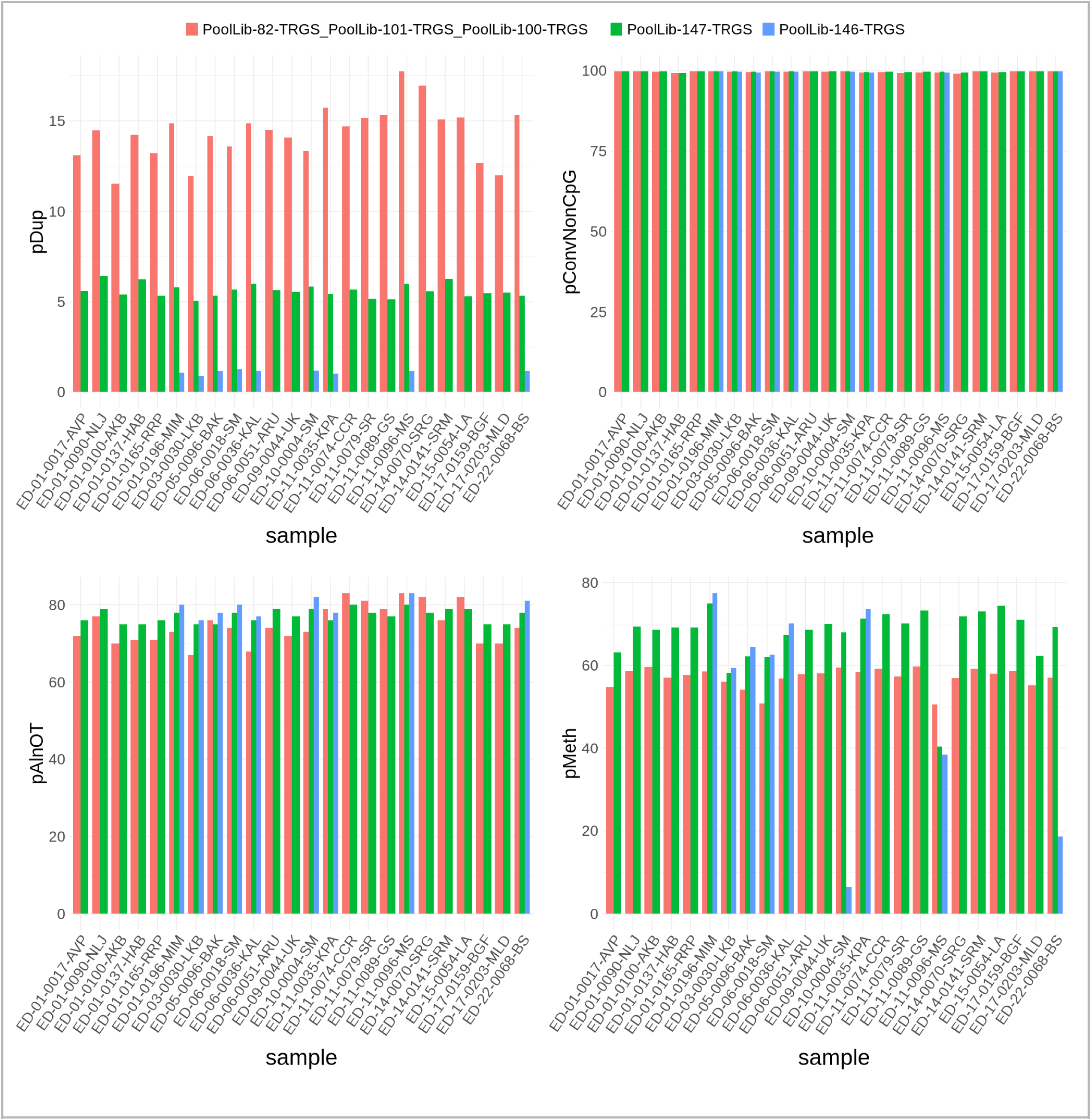
Reproducibility when the same sample was run in different capture pools

**Figure 4.3b:**
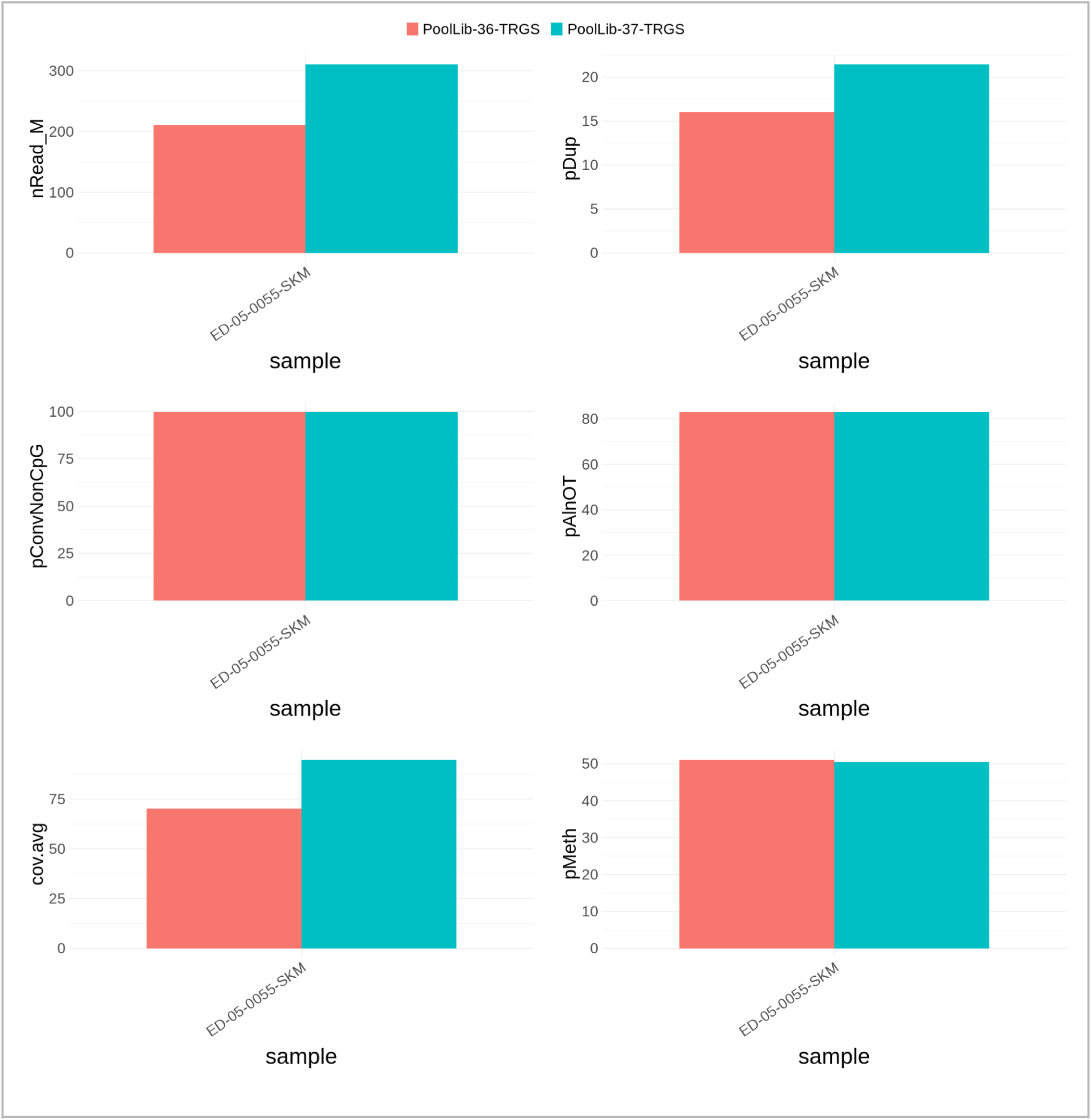
Reproducibility when the same sample ED-05-0055-SKM was run in different capture pools.

**Figure 5.1:**
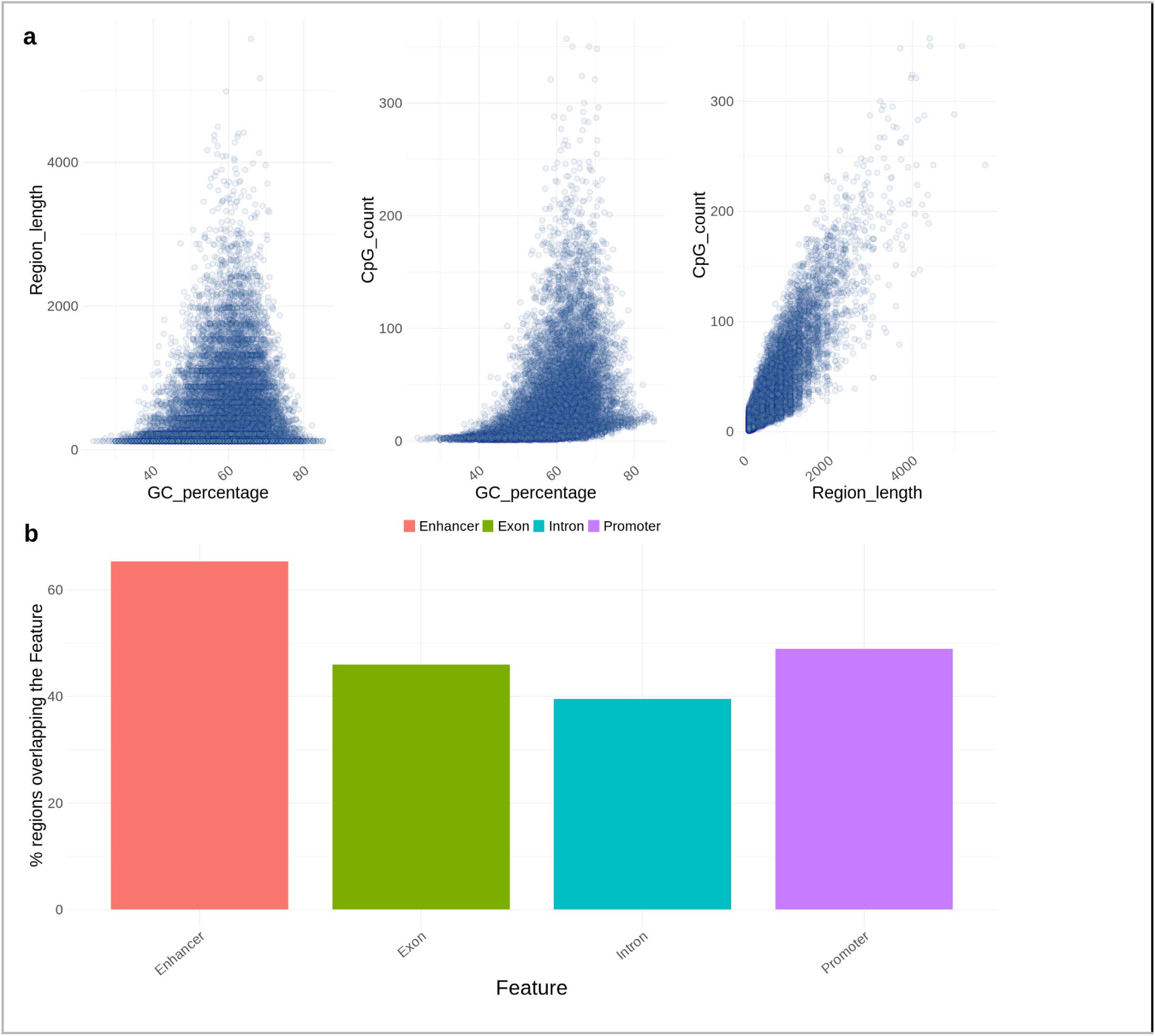
(a) Relationships between key regional features: region length versus GC content (%GC), %GC versus the number of CpG sites, and region length versus the number of CpG sites. (b) Percentage of regions overlapping with genomic annotations, including promoters, enhancers, introns, and exons.

**Figure 6.1:**
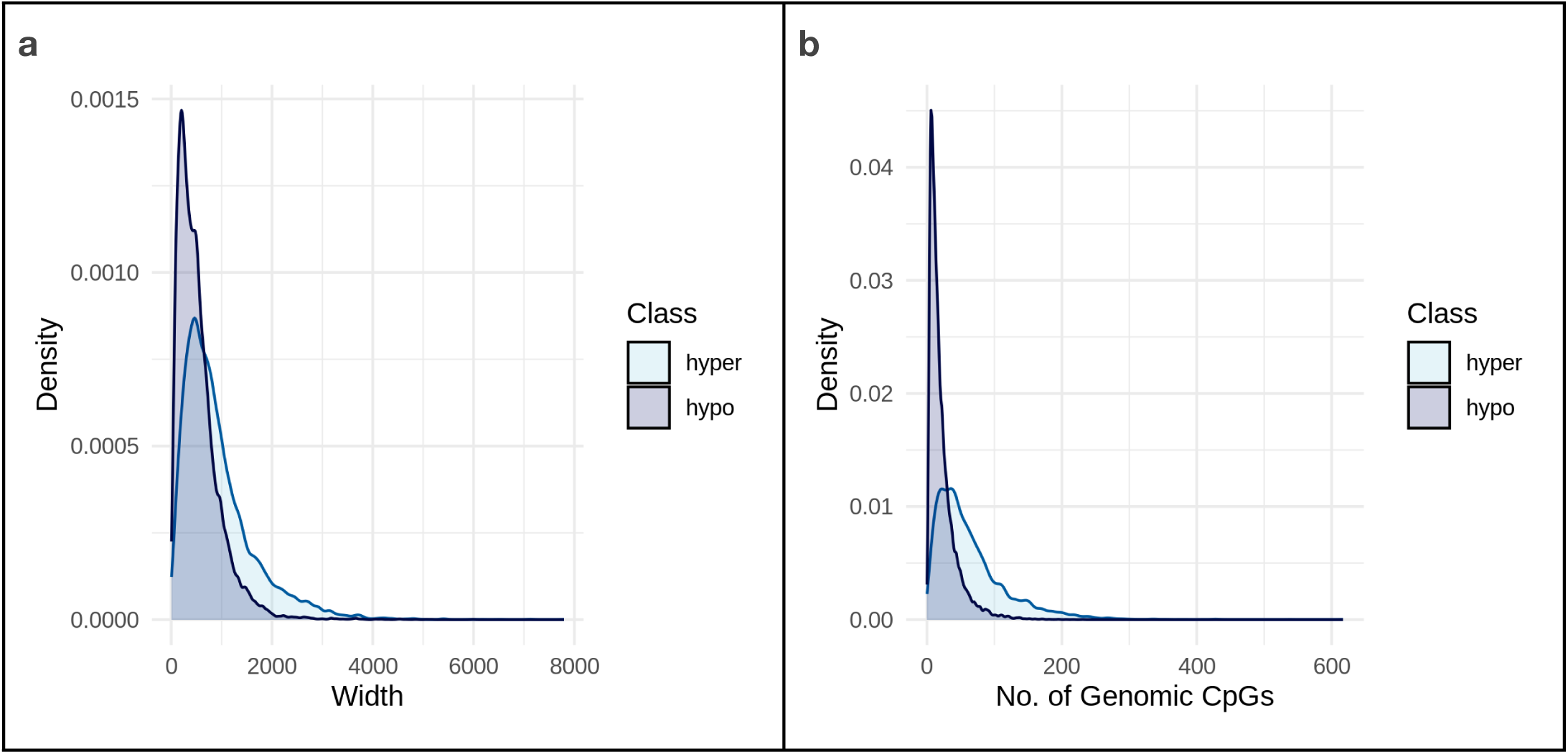
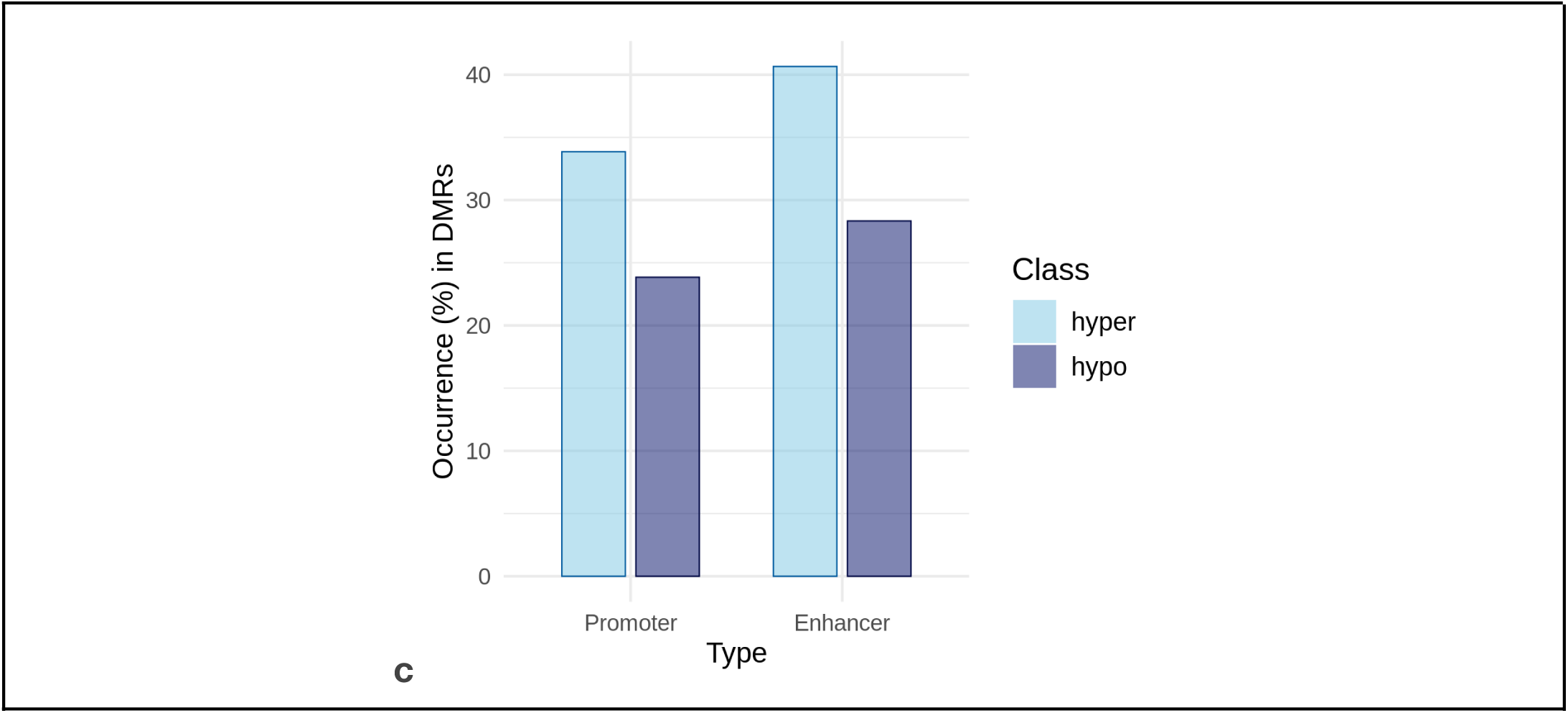
Density plots illustrating the distribution of hypermethylated and hypomethylated differentially methylated regions (DMRs) based on **(a)** their width and **(b)** the number of genomic CpGs covered. **(c)** Bar plot displaying the percentage of occurrences in hypermethylated and hypomethylated DMRs for both promoter and enhancer regions.

### 5. Characteristics of regions selected via ML

A total of 16,937 regions were identified through machine learning (ML) selection, with region lengths ranging from 121 to 5,719 base pairs. These regions were annotated using the csaw [1] package, which provided detailed information on overlaps with RefSeq promoters, introns, and exons. Additionally, the regions were evaluated for overlap with ENCODE enhancers, with 65% of the regions showing enhancer overlap, the highest among the annotated features. The GC content of the regions varied widely, ranging from 24% to 85%, reflecting the diverse genomic contexts represented within the selected regions. These annotations highlight the genomic and regulatory features associated with the regions prioritized by ML.

The percentage of the full Twist Methylome of 551,803 regions overlapping with various genomic features, such as promoters, exons, introns, and enhancers, differs from the overlap observed for the selected 16,937 regions. Notably, the proportion of regions overlapping with enhancers decreases from 65% in the 16,937 regions to 45.8% in the complete 551,803 dataset.

### 6. Public methylation data analysis

#### Datasets Utilized in This Study

This study primarily utilized data generated using <u>Illumina HumanMethylation 450K</u> BeadChips, an assay that interrogates approximately 485,547 CpG sites across the human genome. Widely employed in methylation studies, this platform has been integral to both academic and industrial research, with many datasets made available through public repositories as supplementary material.

#### The TCGA Dataset

The Cancer Genome Atlas (TCGA) (https://www.cancer.gov/tcga ) offers a standardized database of methylation samples, addressing the limitations of GEO data. TCGA employs uniform protocols for sample collection, treatment, and normalization, with beta values generated using the SeSAMe normalization technique [2]. This standardization facilitates reliable comparative analyses and ensures traceability for individual samples. (Note: We primarily used data from Illumina 450K, except in the case of Ovarian cancer, where we utilized Illumina 27K due to unavailability of samples from 450K. )

#### The EWAS Dataset

The EWAS dataset, maintained by the National Genomics Data Center, incorporates samples from both TCGA and GEO datasets. For this study, TCGA cancer samples were excluded, and only GEO-derived cancer samples were included for analysis. EWAS also provides valuable methylation data for various tissues and healthy blood samples, which were leveraged as supplementary datasets. The dataset applies Gaussian Mixture Quantile Normalization (GMQN), complementing TCGA and GEO data. Additional details about the EWAS dataset are provided in the following table.

There are a few cancer types for which we do not have enough either cancer samples or healthy normal samples or both. So, we augment the TCGA and the EWAS dataset with comparable datasets from the GEO.

#### The GEO Test Dataset

The Gene Expression Omnibus (GEO) serves as a key repository for data from published academic studies, including methylation datasets. However, challenges with GEO data include variability in enzymatic/chemical treatments and normalization methods across studies. Such inconsistencies complicate pooling cancer or healthy samples for analysis. While GEO data is suboptimal for identifying cancer-specific methylation patterns, it serves as a valuable source for validation. Specific GEO datasets used in this study are detailed in the following table and used to augment cancer types with low or missing samples from above. These datasets are as follows:

● CESC:

○ TCGA Normals supplement - <u>GSE211668</u> (UK, n=30)
○ EWAS Normal supplement - <u>GSE143752</u> (Canadian, n=54)
● OV:

○ TCGA Cancer - <u>GSE133556</u> (American, n=99), <u>GSE65820</u> (Australian, n=114), <u>GSE72021</u> (Norwegian, n=221)
○ TCGA Healthy - <u>GSE133556</u> (American, n=12), <u>GSE65820</u> (Australian, n=7)
○ EWAS Healthy Tissue - <u>GSE74845</u> (Norwegian, n=52)
● STAD:

○ TCGA Healthy - <u>GSE99553</u> (Korean, n = 84), <u>GSE72872</u> (Australian, n = 82)
● UCEC & UCS:

○ EWAS Healthy Tissue - <u>GSE90060</u> (Estonian, n=68)
● PAAD:

○ PAAD Healthy Tissue - <u>GSE74071</u> (Taiwan, n = 7)
● GEO Healthy Blood:

○ <u>GSE40279</u> (American, n = 636)

#### Seed Probe Selection

To address the high dimensionality of methylation data, a statistical selection approach was employed to identify probes with strong cancer vs. healthy methylation signatures. Probes were filtered based on beta-value distribution and a log2 fold-change threshold, resulting in 20,854 hypermethylated and 60,804 hypomethylated seed probes. These seed probes, indicative of consistent regional methylation trends, were designed to narrow the search space for downstream analyses and facilitate targeted sequencing. In brief for each probe we performed 4 comparisons whose details are given below.

- specificCancer: If the probe shows differential methylation in a given cancer type (TCGA) vs specific tissue type adjacent normals
- pooledAdjNormals: If the probes continue to show a differential trend in a given cancer type compared to the entire TCGA normal cohort.
- healthyBlood: If the probe is differentially methylated in a cancer type compared to methylation in a blood sample cohort.
- healthyTissue: If a probe is differentially methylated in a given cancer type with respect to an independent cohort of healthy tissue samples (eg. lung cancer vs healthy lung tissue)

Using the parameters outlined above, the following cutoffs were determined for each cancer type. Applying these cutoffs, we identified the numbers of hypermethylated and hypomethylated seed probes which are summarized in Supplementary Table 5.

#### Differentially Methylated Regions (DMRs)

To identify DMRs for a given cancer type, we began by selecting seed probes specific to the cancer type of interest. These seed probes were expanded to include all probes within ±2.5 kb of their genomic locations. The beta values for these selected probes were obtained from TCGA cancer samples and corresponding adjacent normal tissues (or GEO normals where adjacent normals were unavailable). The data was processed using two widely cited differential methylation pipelines: Bumphunter 1.34.0, [3]) and DMRcate version 2.6.0; [4]. Both methods yielded approximately 60% overlap in predicted bases for LUAD. DMRcate was ultimately selected for downstream analysis due to its higher precision, as demonstrated in benchmark studies. Genome browser visualizations further confirmed that DMRcate-predicted regions were cancer-specific.

To refine our approach, we tested DMRcate with the full set of 409k high-quality probes from the Illumina 450K array, as well as with the seed probes plus their ±2.5 kb flanking regions. For COAD, the comparison of top-ranked probes between the two approaches revealed significant overlap (16 out of the top 20 probes were common). This suggests that seed probes, often representing CpG sites hypermethylated in cancer (or hypomethylated in adjacent normal tissue), effectively capture the broader methylation trends of their genomic neighborhoods. Leveraging seed probes and their flanks helps reduce the search space while maintaining accuracy in identifying significant regions.

#### DMR Identification Workflow

Based on these findings, the following steps were adopted:

1. Select seed hypermethylated probes for a specific cancer type (e.g., BRCA).
2. Identify all other probes within ±2.5 Kb of the seed probes.
3. Process the pool of probes through DMRcate to identify DMRs.
4. Repeat the process for seed hypomethylated probes.

#### DMRcate Parameters

● Minimum DMR length: 300 bp
● Minimum number of 450K probes per DMR: 3
● Lambda (distance threshold for assigning CpGs to different DMRs): 500 bp
● C (scaling factor based on 1 SD of the beta peak): 5 (resulting in lambda/C = 100 bp)
● False discovery rate (FDR): 0.05

The lambda and C values were adapted from a benchmarking study on methylation arrays [5] . The resulting DMRs, stratified into hypermethylated and hypomethylated regions, are provided in Supplementary Table 5. Across multiple cancer types, 10,494 unique hypermethylated and 7,399 unique hypomethylated DMRs were predicted.

#### Annotations for Each DMR

To enrich the analysis, we added the following annotations (beyond those generated by DMRcate):

● IDs of probes within the DMR.
● Number of genomic CpG sites within the DMR (noting the contrast between the 450K array probes and ∼29 million CpGs in the human genome).
● Mean and maximum genomic CpGs when the DMR is divided into 100 bp bins (useful for designing 120-bp hybridization probes).
● Number of UCSC RefSeq promoters, ENCODE enhancers, and DNase hypersensitivity clusters overlapping each DMR.

● Overlap of DMR-associated genes/probes with those identified via curated literature reviews.

---

**Table 5.1.**
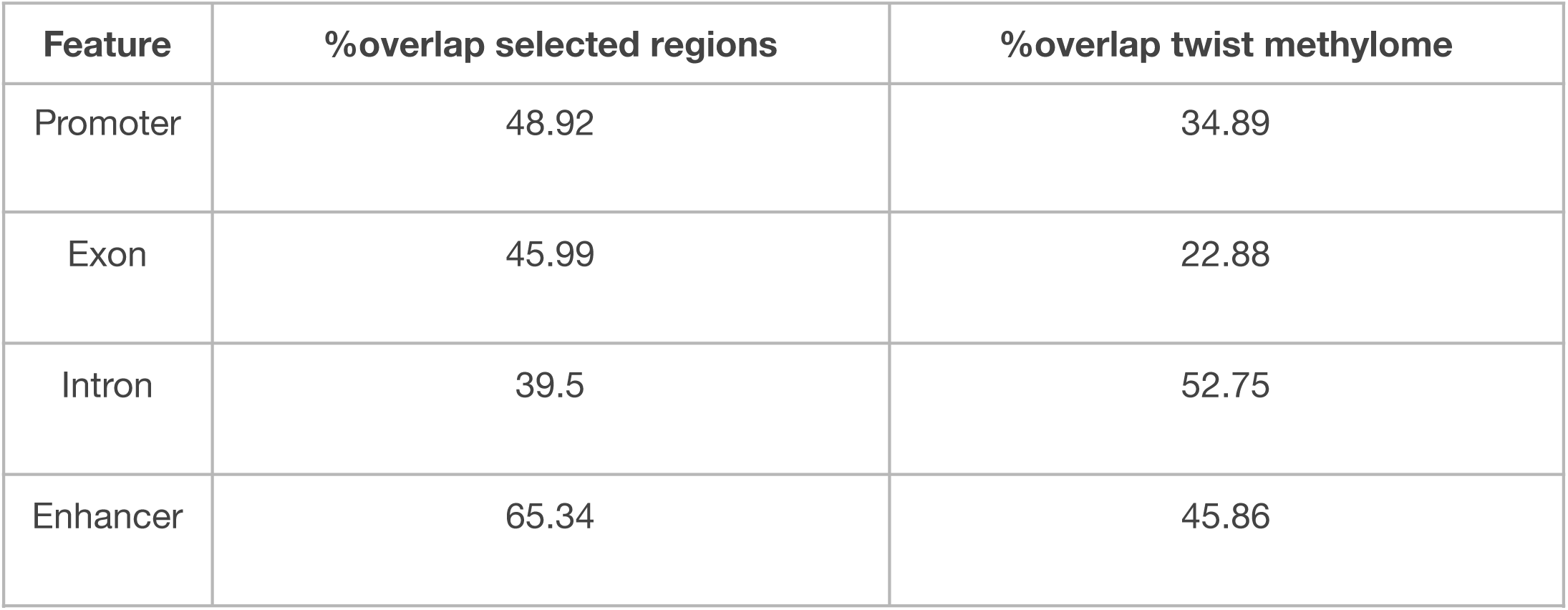
Percentage of genomic features overlapping the entire twist methylome panel compared to regions selected by the ML analysis.

| Feature | %overlap selected regions | %overlap twist methylome |
| --- | --- | --- |
| Promoter | 48.92 | 34.89 |
| Exon | 45.99 | 22.88 |
| Intron | 39.5 | 52.75 |
| Enhancer | 65.34 | 45.86 |

**Table 6.1.** Breakdown of TCGA samples for analysis, categorized by cancer type. The table includes the full name of each cancer type and the distribution of cancer and healthy samples, grouped by assay type (450K and 27K arrays).

| Cancer Types | Full Name | Cancer Samples - 450K | Healthy Samples - 450K | Cancer Samples - 27K | Healthy Samples - 27K |
| --- | --- | --- | --- | --- | --- |
| BRCA | Breast Cancer | 793 | 97 | 314 | 27 |
| CESC | Cervical Squamous Cell Carcinoma | 307 | 3 | 0 | 0 |
| COAD | Colon Adenocarcinoma | 312 | 38 | 166 | 37 |
| ESCA | Esophageal Carcinoma | 185 | 16 | 0 | 0 |
| LUAD | Lung Adenocarcinoma | 473 | 32 | 126 | 24 |
| LUSC | Lung Squamous Cell Carcinoma | 370 | 42 | 134 | 27 |
| OV | Ovarian Cancer | 10 | 0 | 601 | 0 |
| READ | Rectum Adenocarcinoma | 98 | 7 | 68 | 5 |
| STAD | Stomach Adenocarcinoma | 395 | 2 | 48 | 25 |
| UCEC | Uterine Corpus Endometrial Carcinoma | 438 | 46 | 117 | 1 |
| UCS | Uterine Carcinoma | 57 | 0 | 0 | 0 |
| LIHC | Liver Hepatocellular Carcinoma | 377 | 50 | 0 | 0 |
| PAAD | Pancreatic Adenocarcinoma | 184 | 10 | 0 | 0 |

**Table 6.2.** Summary of samples extracted from EWAS for analysis, categorized by cancer type. The table includes the full name of each cancer type, the total number of cancer samples, and the breakdown of samples by source: tissue samples (450K), TCGA samples, and GEO samples.

| Cancer Types | Full Name | Cancer Samples | Tissue Samples 450K | TCGA Samples | GEO Samples |
| --- | --- | --- | --- | --- | --- |
| BRCA | Breast Cancer | 939 | 301 | 297 | 641 |
| CESC | Cervical Squamous Cell Carcinoma | 225 | 0 | 198 | 26 |
| COAD | Colon Adenocarcinoma | 229 | 239 | 155 | 73 |
| ESCA | Esophageal Carcinoma | 296 | 126 | 190 | 105 |
| HNSC | Head and Neck Squamous Cell Carcinoma | 471 | 0 | 411 | 59 |
| LUAD | Lung Adenocarcinoma | 339 | 301 | 166 | 172 |
| LUSC | Lung Squamous Cell Carcinoma | 323 | 301 | 260 | 62 |
| OV | Ovarian Cancer | 0 | 0 | 0 | 0 |
| READ | Rectum Adenocarcinoma | 0 | 0 | 0 | 0 |
| STAD | Stomach Adenocarcinoma | 482 | 146 | 393 | 88 |
| UCEC | Uterine Corpus Endometrial Carcinoma | 263 | 0 | 225 | 37 |
| UCS | Uterine Carcinoma | 117 | 0 | 38 | 78 |
| LIHC | Liver Hepatocellular Carcinoma | 306 | 300 | 89 | 216 |
| PAAD | Pancreatic Adenocarcinoma | 179 | 75 | 178 | 0 |

**Table 6.3.** LogFC and p-value cut-offs used to select seed probes.

| Name | Type | logFC cut-off | Pval cut-off |
| --- | --- | --- | --- |
| specificCancer | Hyper | >90th percentile | 0.01 |
| pooledAdjNormals | Hyper | >70th percentile | 0.05 |
| healthyblood | Hyper | >90th percentile | 0.05 |
| healthyTissue | Hyper | >70th percentile | 0.05 |
| specificCancer | Hypo | <10th percentile | 0.01 |
| pooledAdjNormals | Hypo | <10th percentile | 0.05 |
| healthyblood | Hypo | <10th percentile | 0.05 |
| healthyTissue | Hypo | <10th percentile | 0.05 |

**Supplementary Figure 1:**
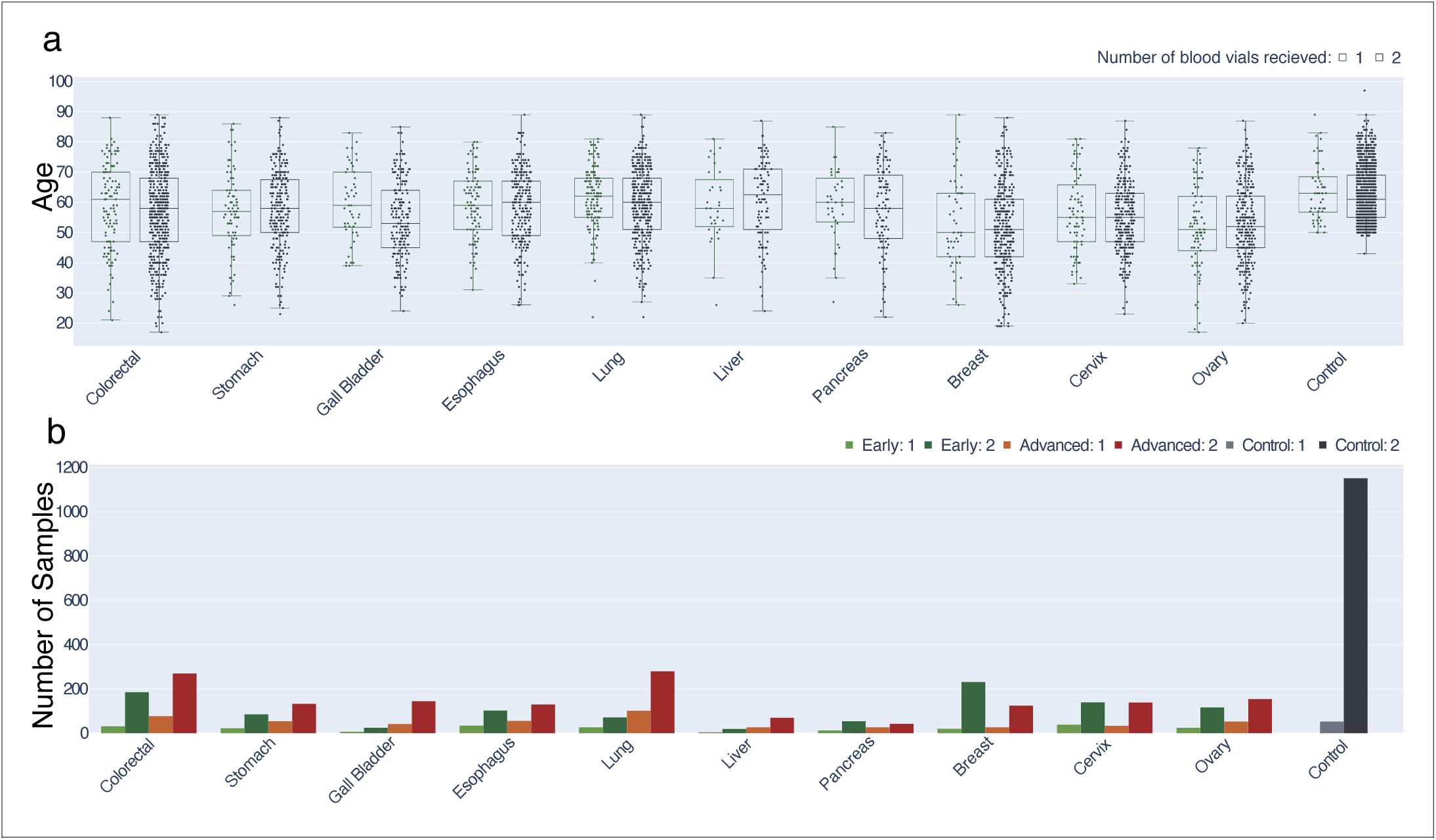
Age distribution across cancer types and controls, and sample distribution by cancer stages. **(a)** Variation in age across different cancer types and controls. The x-axis represents the cancer types or control groups, while the y-axis displays the age of the samples. For each cancer type or control group, two distributions are shown, stratified by the number of blood tubes collected, allowing for a comparison of age distributions within these subgroups. **(b)** Distribution of samples across various cancer types and their respective stages, providing insight into the dataset composition by cancer stage.

**Supplementary Figure 2:**
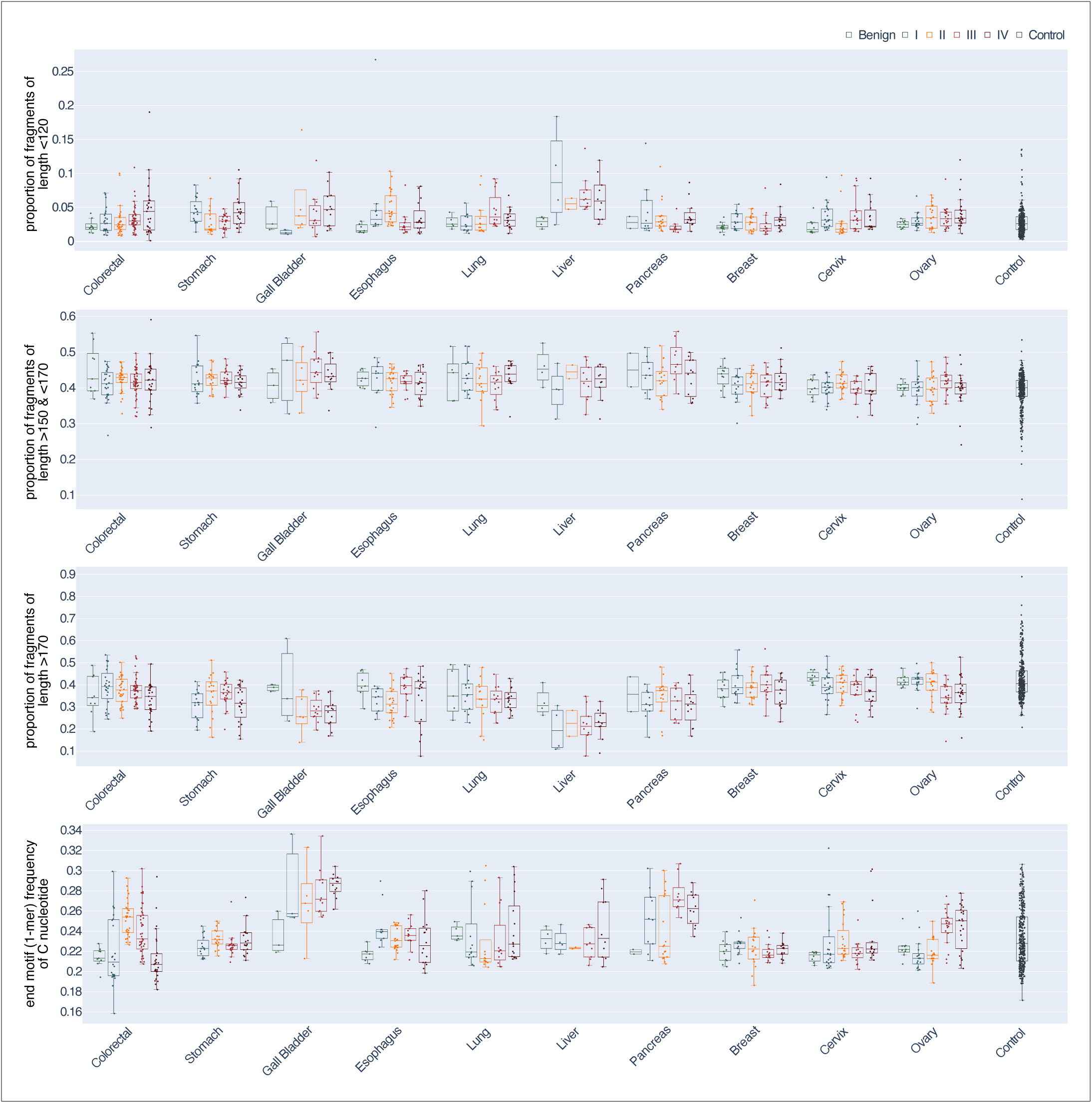
Variation in fragment length motifs across cancer types and control groups. Distribution of fragment lengths across different cancer types and control groups. The y-axis represents the proportion of fragments categorized by length: <120 bp, between 150 and 170 bp, and >170 bp. Additionally, the frequency of nucleotide end motifs (1-mers) is shown for each group, providing insights into the distribution of these motifs across the samples.

**Supplementary Figure 3:**
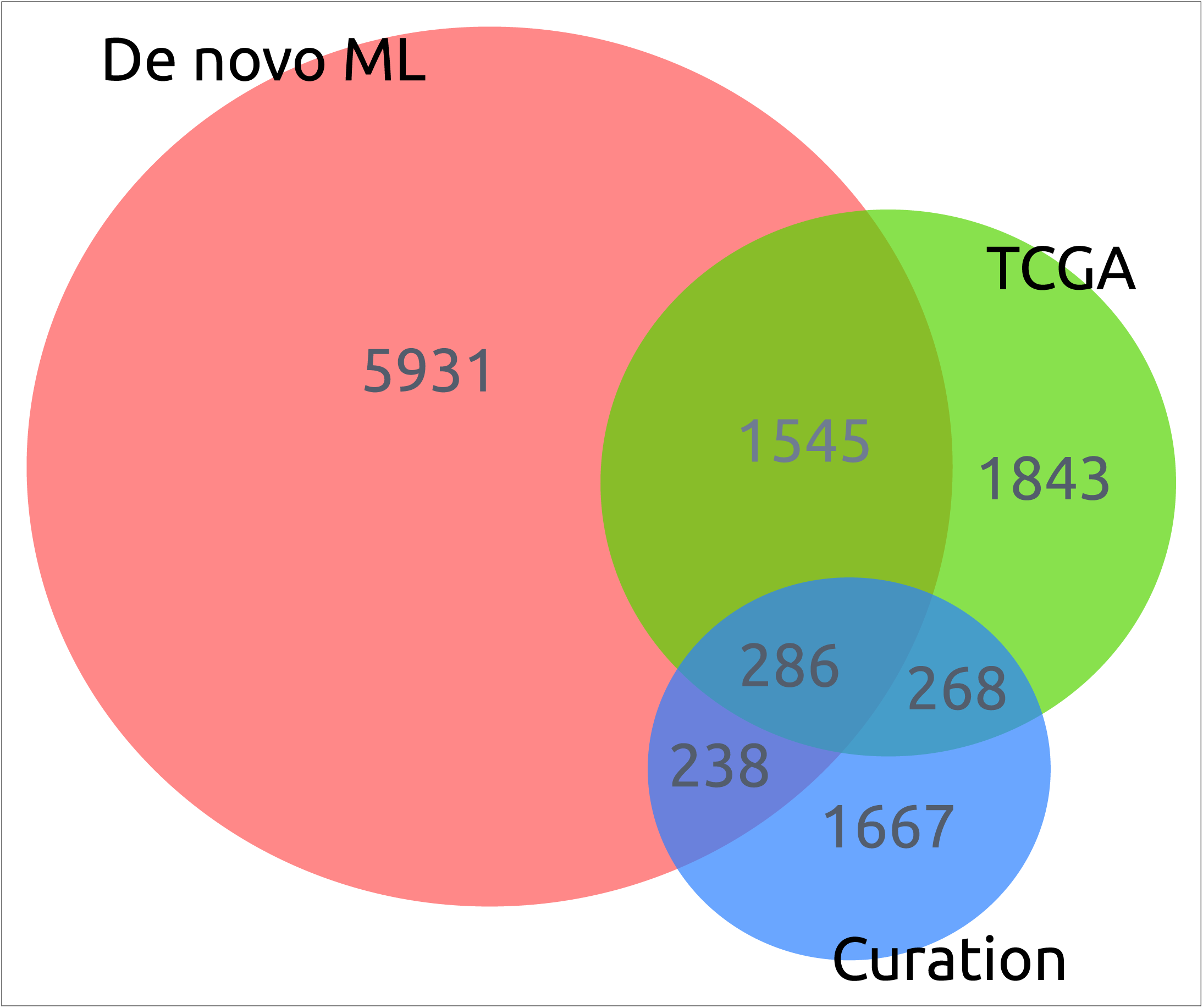
Overlap of Genomic Regions from De Novo ML-Selected, TCGA-Derived, and Curated Datasets. Intersection of genomic regions identified from three sources: the de novo machine learning (ML)-selected regions (7,460 regions), TCGA-derived regions (3,942 regions), and curated regions (2,459 regions), thus showing the overlap and distinct genomic regions identified by each method.

**Supplementary Figure 4:**
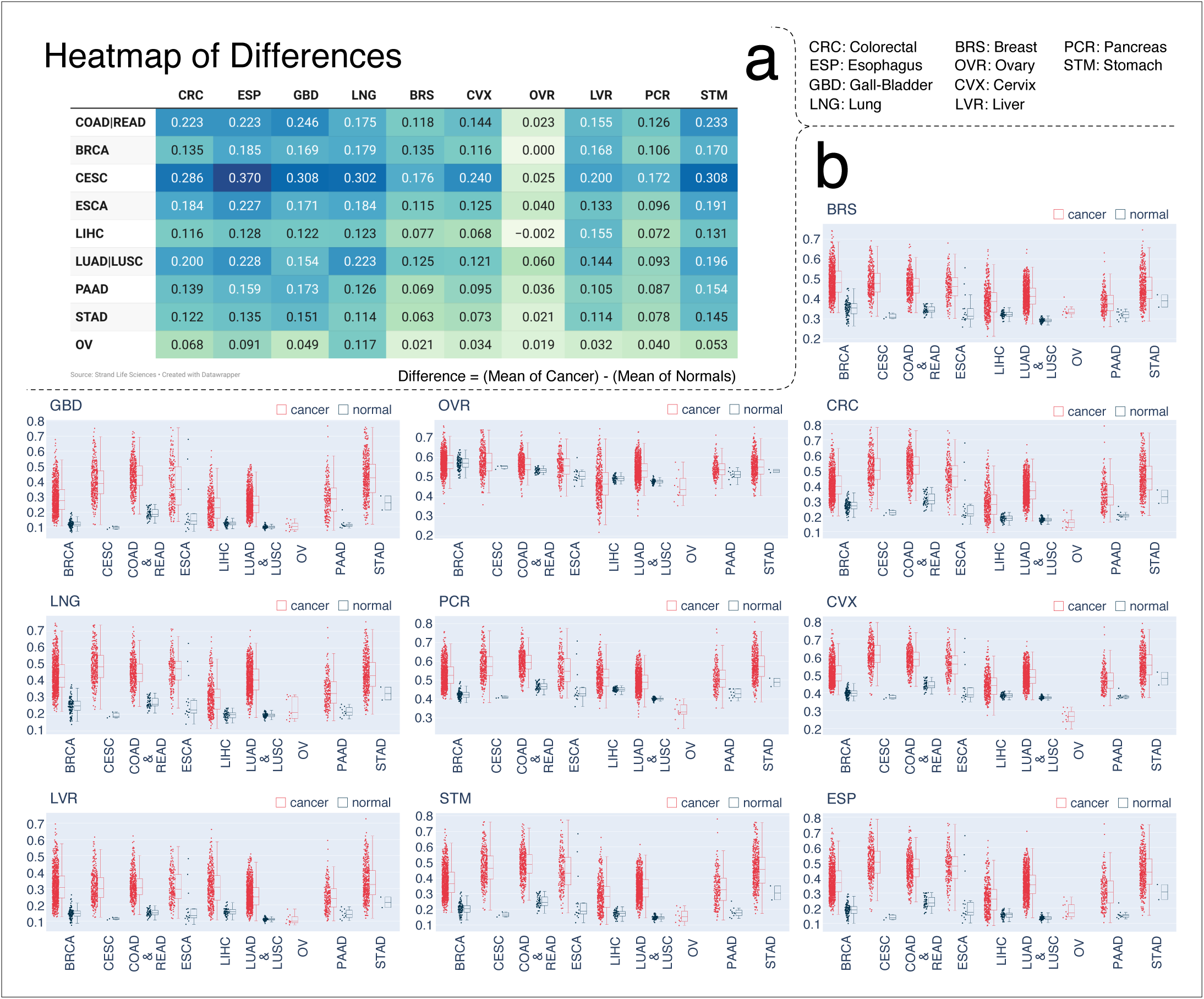
Methylation Distribution of Cancer and Adjacent Normal Tissues Across TCGA Cancer Types. **(a)** Heatmap where columns represents TCGA cancer samples, and rows correspond to regions showing differential methylation scores between each cancer type and its relevant control. Each cell reflects the difference in average methylation values between cancer tissue and adjacent normal tissue across all regions in the TCGA dataset. This visualization highlights the differential methylation patterns observed in cancer samples compared to adjacent normal tissues. **(b)** The x-axis represents different TCGA cancer types. For each cancer type, two plots are presented: one for the cancer tissue and one for the adjacent normal tissue. These plots display the distribution of mean methylation values for each group, highlighting the differences in methylation between cancer and adjacent normal tissues across the selected regions from the models.

